# Estimating the number of SARS-CoV-2 infections in the United States

**DOI:** 10.1101/2020.04.13.20064519

**Authors:** Dayton G. Thorpe, Kelsey Lyberger

## Abstract

We apply a model developed by The COVID-19 Response Team [S. Flaxman, S. Mishra, A. Gandy, *et al*., “Estimating the number of infections and the impact of non-pharmaceutical interventions on COVID-19 in 11 European countries,” tech. rep., Imperial College London, 2020.] to estimate the total number of SARS-CoV-2 infections in the United States. Across the United States we estimate as of April 18, 2020 the fraction of the population infected was 4.6% [3.6%, 5.8%], 21 times the portion of the population with a positive test result. Excluding New York state, which we estimate accounts for over half of infections in the United States, we estimate an infection rate of 2.3% [2.1%, 2.8%].

We include the timing of each state’s implementation of interventions including encouraging social distancing, closing schools, banning public events, and a lockdown / stay-at-home order. We assume fatalities are reported correctly and infer the number and timing of infections based on the infection fatality rate measured in populations that were tested universally for SARS-CoV-2. Underreporting of deaths would drive our estimates to be too low. Reporting of deaths on the wrong day could drive errors in either direction. This model does not include effects of herd immunity; in states where the estimated infection rate is very high - namely, New York - our estimates may be too high.

## I. Introduction

The first case of SARS-CoV-2, the virus that causes COVID-19, diagnosed in the United States was discovered on January 21, 2020 [1]. Since then, limited availability of SARS-CoV-2 tests has interfered with efforts to track the spread of the virus. As of April 18, 2020, according to Johns Hopkins University there were 732,200 reported cases in the United States [2]. The true number of cases could be far higher.

Europe has faced similar constraints on SARS-CoV-2 testing. The COVID-19 Response Team (CRT) at Imperial College London has responded to this challenge, in part, by developing a Bayesian hierarchical model to estimate the true number of infections and the impact of policies intended to slow the epidemic [3]. In this paper, we apply the CRT model to individual states in the United States, plus Puerto Rico and Washington, D.C. We remove states with fewer than 10 reported fatalities. We also reproduce the CRT estimates for 11 European countries with data through April 18, 2020.

We briefly summarize the CRT model here, but refer to [3] for full details. The model takes three inputs from each modeled population: 1) the age distribution, 2) the time series of known COVID-19 fatalities, and 3) the start dates of four mitigation policies (encouraging social distancing, closing schools and universities, banning public events, and a lockdown / stay-at-home order). It also relies on four values estimated from prior research: 1) the infection fatality ratio (IFR) by age, 2) the serial interval distribution, 3) the infection-to-onset distribution, and 4) the onset-to-death distribution. These values are measured from several cases in which all members of a small population at high risk of infection were tested and observed, whether or not they showed symptoms. They are not extracted from population data based on testing people who are already symptomatic. We discuss the IFR further in section III.

There are several possible sources of error we do not account for. First, we assume COVID-19 fatalities are reported correctly. If they are underreported, the infection estimates here will be underestimates. If they are reported on the wrong day, the estimates could be distorted in either direction. Second, the model extrapolates exponential growth rates without reference to the size of the population. If the estimated attack rate approaches the level of herd immunity, the model likely overestimates the total number of infections. Third, we assume the IFR is known exactly. Propagating the error from the IFR would increase the width of our confidence intervals.

The model estimates the baseline reproduction number of the virus without intervention, R_0, and the impact of the considered interventions. It fits one value for each of these variables across all populations simultaneously. The impact of any given intervention is the same for all populations. Most of the interventions were implemented in Europe before the United States. Therefore, the model estimates the impact of these interventions mostly based on the trajectory of the epidemic in Europe. Using the number of fatalities as of a given time, we can infer the number of infections in the past, and extrapolate them to the present using the fitted reproduction numbers under different policy regimes. The model does not use the reported number of infections.

This approach differs from the CRT’s model in [4], which is a detailed SIR model. Instead, this model more closely matches that of the UW IHME [5], which forecasts exponential growth rates after different policies are implemented based on historical growth rates in countries and states that previously implemented similar policies.

## II. Results

As in Europe, we estimate infections in the United States are much higher than reported infections. As of April 18, 2020 we estimate the attack rate in the United States was 4.6% [3.6%, 5.8%], 21 times the reported infections of 732,200. The difference between estimated and reported infections varies substantially by state. In New York, there are an estimated 7,600,000 infections, 32 times the reported value of 237,000. Whereas, in Utah, our estimate of infections is only 3.0 times the reported value.

In Figure 1 and Table 1 we report the estimated fraction of the population infected in each state. The highest estimate is in New York, at 39.8% [22.8%, 56.9%], followed by New Jersey at 19.7% [12.0%, 33.4%].

**Table 1.** Fraction of the population infected for each state as of April 18, 2020, estimated from our model.

| Estimates as of April 18, 2020 |  |  |  |
| --- | --- | --- | --- |
| State | Infections | % of population infected | 95% confidence interval |
| Alabama | 40,880 | 0.84% | [0.46%, 1.40%] |
| Arizona | 43,054 | 0.62% | [0.36%, 0.97%] |
| Arkansas | 13,886 | 0.46% | [0.12%, 1.48%] |
| California | 329,652 | 0.84% | [0.53%, 1.21%] |
| Colorado | 96,621 | 1.75% | [1.09%, 2.80%] |
| Connecticut | 427,256 | 11.93% | [7.17%, 17.72%] |
| Delaware | 17,003 | 1.79% | [0.88%, 3.68%] |
| DC | 34,494 | 5.04% | [2.46%, 9.12%] |
| Florida | 204,287 | 0.99% | [0.62%, 1.64%] |
| Georgia | 204,632 | 1.99% | [1.23%, 2.85%] |
| Idaho | 11,612 | 0.69% | [0.25%, 1.62%] |
| Illinois | 507,065 | 3.95% | [2.23%, 6.94%] |
| Indiana | 156,872 | 2.36% | [1.45%, 3.91%] |
| Iowa | 52,128 | 1.66% | [0.56%, 3.59%] |
| Kansas | 21,067 | 0.72% | [0.32%, 1.42%] |
| Kentucky | 35,635 | 0.80% | [0.40%, 1.66%] |
| Louisiana | 306,545 | 6.57% | [4.24%, 10.61%] |
| Maine | 6,620 | 0.50% | [0.18%, 1.07%] |
| Maryland | 164,973 | 2.75% | [1.62%, 4.54%] |
| Massachusetts | 563,979 | 8.26% | [4.58%, 13.65%] |
| Michigan | 673,261 | 6.76% | [4.42%, 10.47%] |
| Minnesota | 34,298 | 0.62% | [0.31%, 1.08%] |
| Mississippi | 45,447 | 1.52% | [0.81%, 2.85%] |
| Missouri | 70,561 | 1.16% | [0.62%, 2.00%] |
| Nebraska | 10,035 | 0.53% | [0.09%, 1.53%] |
| Nevada | 37,955 | 1.30% | [0.75%, 2.15%] |
| New Hampshire | 10,577 | 0.79% | [0.27%, 1.74%] |
| New Jersey | 1,752,179 | 19.73% | [12.00%, 33.42%] |
| New Mexico | 17,326 | 0.83% | [0.34%, 1.49%] |
| New York | 7,616,534 | 38.82% | [22.76%, 56.88%] |
| North Carolina | 61,299 | 0.60% | [0.34%, 1.07%] |
| Ohio | 114,744 | 0.99% | [0.57%, 1.65%] |
| Oklahoma | 28,854 | 0.74% | [0.41%, 1.21%] |
| Oregon | 13,026 | 0.32% | [0.18%, 0.55%] |
| Pennsylvania | 383,651 | 3.00% | [1.84%, 4.90%] |
| Puerto Rico | 11,767 | 0.35% | [0.16%, 0.75%] |
| Rhode Island | 48,463 | 4.59% | [2.15%, 9.37%] |
| South Carolina | 31,708 | 0.64% | [0.33%, 1.21%] |
| Tennessee | 30,408 | 0.46% | [0.26%, 0.73%] |
| Texas | 179,211 | 0.64% | [0.41%, 1.01%] |
| Utah | 8,753 | 0.29% | [0.10%, 0.64%] |
| Vermont | 5,454 | 0.87% | [0.33%, 1.79%] |
| Virginia | 89,638 | 1.07% | [0.63%, 1.75%] |
| Washington | 95,882 | 1.31% | [0.96%, 1.78%] |
| West Virginia | 5,150 | 0.28% | [0.06%, 0.87%] |
| Wisconsin | 48,269 | 0.84% | [0.47%, 1.43%] |

**Figure 1.**
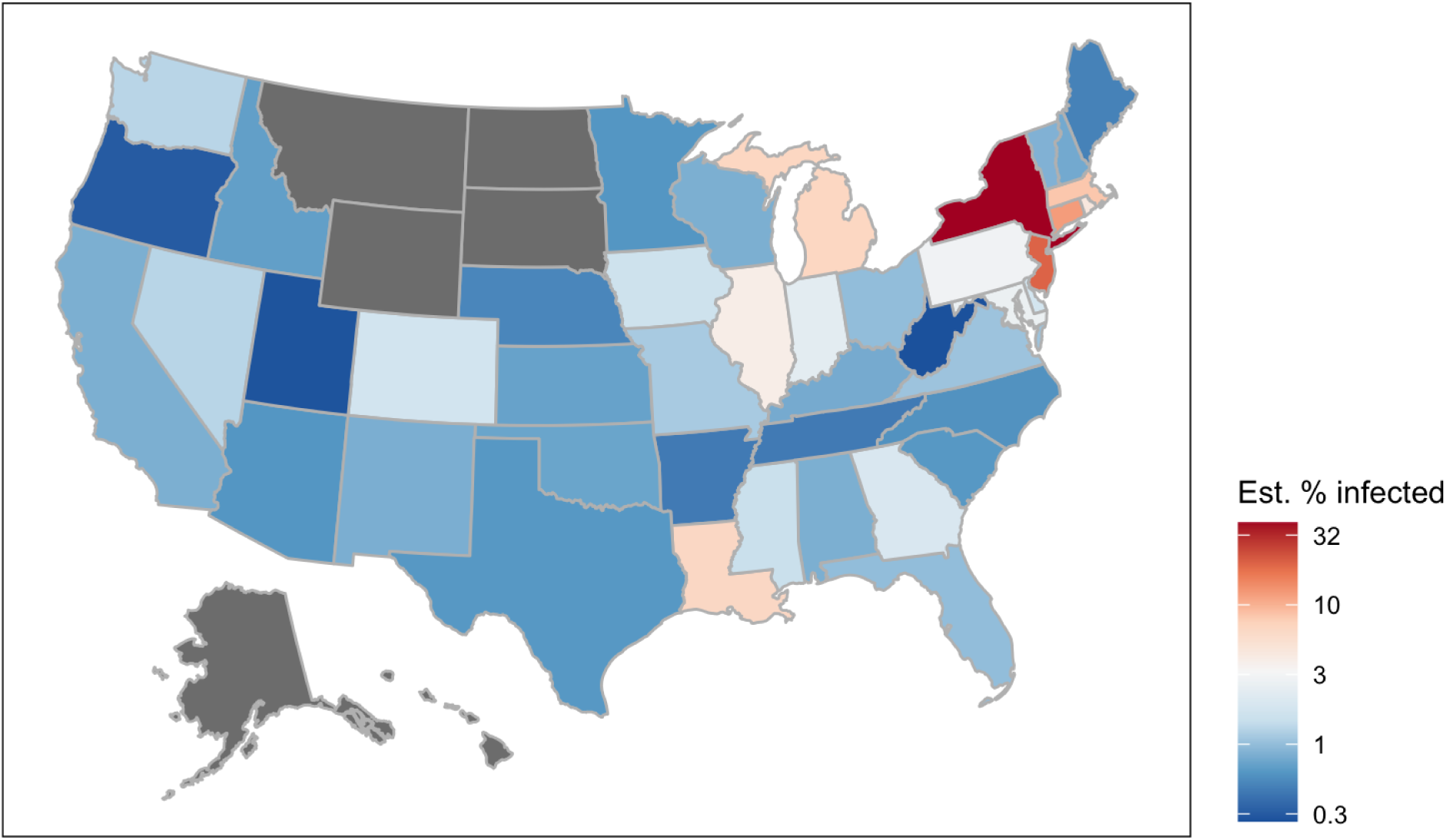
Log-transformed percent of the population infected for each state as of April 18, 2020, estimated from our model.

In Table 2, we compare the weighted average attack rate in the United States to the same 11 European countries as in [3].

**Table 2.** Fraction of the population infected for each of 11 European countries and the United States as of April 18, 2020, estimated from our model. *The fraction of the population infected for the United States is the population-weighted average of the states we are able to model. Those states sum to 98% of the US population.

|  | **Estimates as of April 18, 2020** |  |  |
| --- | --- | --- | --- |
| Country | **Infections** | **% of population infected** | **95% confidence interval** |
| Austria | 91,762 | 1.02% | [0.66%, 1.49%] |
| Belgium | 2,191,275 | 18.91% | [11.61%, 31.31%] |
| Denmark | 69,074 | 1.19% | [0.71%, 1.85%] |
| France | 4,162,245 | 6.38% | [4.32%, 9.24%] |
| Germany | 1,063,715 | 1.27% | [0.86%, 1.90%] |
| Italy | 3,305,547 | 5.47% | [4.06%, 7.21%] |
| Norway | 35,104 | 0.65% | [0.36%, 1.09%] |
| Spain | 4,257,641 | 9.11% | [6.22%, 12.85%] |
| Sweden | 1,560,858 | 15.46% | [9.49%, 23.72%] |
| Switzerland | 235,369 | 2.72% | [1.77%, 4.03%] |
| United Kingdom | 4,748,818 | 7.00% | [4.81%, 9.78%] |
| United States* |  | 4.57% | [3.55%, 5.76%] |

In each population, the model infers the time series of infections that most likely gave rise to the observed time series of fatalities. In the appendix, in Figure A1 through Figure A56, we show the estimated and reported time series of infections and fatalities. Model parameters are fit jointly across all populations. The first panel in each of these figures shows how well the model reproduces the observed fatalities. The gap between estimated and reported fatalities can be interpreted, in part, as a measure of how well the model reproduces the epidemic in each population.

## III. Methods

We retrieved dates for policy interventions as follows. We took lockdown dates from the *New York Times* rounded to the nearest day (lockdowns going into effect past noon are recorded as the next day) [6]. School closure dates are from [7]. Following [3], if a school closure began on a Monday, we set the date of the school closure to the previous Saturday. We set social distancing and the ban of public events to Mar 16, 2020 for all states, when federal guidelines were put into place [8]. See the dates we assign for each region in each state in the Appendix (Table A1). Age distributions for each state come from the US Census Bureau [9]. The time series of reported fatalities and cases in the United States comes from the Johns Hopkins dashboard [2]. The time series of reported fatalities and cases in the 11 European countries comes from the European Centre of Disease Control (ECDC) [10]. We change the name of the country “Georgia” to “Georgia(country)” in the ECDC data set to avoid confusion with the state of Georgia. In the ECDC data we also delete the entries for Puerto Rico, for which we get duplicate data as a US territory. For the 11 European countries, IFR comes from [3]. All other IFR’s are age-adjusted from [11].

All of our code and input data are available at https://drive.google.com/drive/folders/1DDST4e3B875wxsVApJTKLn94seM5IZbx?usp=sharing.

## Data Availability

All of our code and input data are available at the Google Drive link below.

https://drive.google.com/drive/folders/1DDST4e3B875wxsVApJTKLn94seM5IZbx?usp=sharing

## Appendix

**Table A1.** Dates we assign for each suppression measure in each state.

| Country | schools<br>universities | travel<br>restrictions | public events | lockdown | social_distancing_e<br>ncouraged | self_isolating_i<br>f_ill |
| --- | --- | --- | --- | --- | --- | --- |
| Alabama | 3/19/20 | 4/5/20 | 3/16/20 | 4/5/20 | 3/16/20 | 4/5/20 |
| Arizona | 3/14/20 | 4/1/20 | 3/16/20 | 4/1/20 | 3/16/20 | 4/1/20 |
| Arkansas | 3/17/20 | 5/1/20 | 3/16/20 | 5/1/20 | 3/16/20 | 5/1/20 |
| California | 3/19/20 | 3/19/20 | 3/16/20 | 3/19/20 | 3/16/20 | 3/19/20 |
| Colorado | 3/21/20 | 3/26/20 | 3/16/20 | 3/26/20 | 3/16/20 | 3/26/20 |
| Connecticut | 3/17/20 | 3/24/20 | 3/16/20 | 3/24/20 | 3/16/20 | 3/24/20 |
| Delaware | 3/14/20 | 3/24/20 | 3/16/20 | 3/24/20 | 3/16/20 | 3/24/20 |
| DC | 3/14/20 | 4/1/20 | 3/16/20 | 4/1/20 | 3/16/20 | 4/1/20 |
| Florida | 3/14/20 | 4/3/20 | 3/16/20 | 4/3/20 | 3/16/20 | 4/3/20 |
| Georgia | 3/18/20 | 4/3/20 | 3/16/20 | 4/3/20 | 3/16/20 | 4/3/20 |
| Idaho | 3/21/20 | 3/26/20 | 3/16/20 | 3/26/20 | 3/16/20 | 3/26/20 |
| Illinois | 3/17/20 | 3/22/20 | 3/16/20 | 3/22/20 | 3/16/20 | 3/22/20 |
| Indiana | 3/19/20 | 3/25/20 | 3/16/20 | 3/25/20 | 3/16/20 | 3/25/20 |
| Iowa | 3/14/20 | 5/1/20 | 3/16/20 | 5/1/20 | 3/16/20 | 5/1/20 |
| Kansas | 3/18/20 | 3/30/20 | 3/16/20 | 3/30/20 | 3/16/20 | 3/30/20 |
| Kentucky | 3/14/20 | 3/26/20 | 3/16/20 | 3/26/20 | 3/16/20 | 3/26/20 |
| Louisiana | 3/14/20 | 3/24/20 | 3/16/20 | 3/24/20 | 3/16/20 | 3/24/20 |
| Maine | 4/2/20 | 4/2/20 | 3/16/20 | 4/2/20 | 3/16/20 | 4/2/20 |
| Maryland | 3/14/20 | 3/31/20 | 3/16/20 | 3/31/20 | 3/16/20 | 3/31/20 |
| Massachusetts | 3/17/20 | 3/24/20 | 3/16/20 | 3/24/20 | 3/16/20 | 3/24/20 |
| Michigan | 3/14/20 | 3/24/20 | 3/16/20 | 3/24/20 | 3/16/20 | 3/24/20 |
| Minnesota | 3/18/20 | 3/28/20 | 3/16/20 | 3/28/20 | 3/16/20 | 3/28/20 |
| Mississippi | 3/20/20 | 4/4/20 | 3/16/20 | 4/4/20 | 3/16/20 | 4/4/20 |
| Missouri | 3/19/20 | 4/6/20 | 3/16/20 | 4/6/20 | 3/16/20 | 4/6/20 |
| Montana | 3/14/20 | 3/28/20 | 3/16/20 | 3/28/20 | 3/16/20 | 3/28/20 |
| Nebraska | 3/21/20 | 5/1/20 | 3/16/20 | 5/1/20 | 3/16/20 | 5/1/20 |
| Nevada | 3/14/20 | 4/1/20 | 3/16/20 | 4/1/20 | 3/16/20 | 4/1/20 |
| New_Hampshire | 3/14/20 | 3/28/20 | 3/16/20 | 3/28/20 | 3/16/20 | 3/28/20 |
| New_Jersey | 3/18/20 | 3/22/20 | 3/16/20 | 3/22/20 | 3/16/20 | 3/22/20 |
| New_Mexico | 3/14/20 | 3/24/20 | 3/16/20 | 3/24/20 | 3/16/20 | 3/24/20 |
| New_York | 3/18/20 | 3/23/20 | 3/16/20 | 3/23/20 | 3/16/20 | 3/23/20 |
| North_Carolina | 3/14/20 | 3/31/20 | 3/16/20 | 3/31/20 | 3/16/20 | 3/31/20 |
| Ohio | 3/17/20 | 3/24/20 | 3/16/20 | 3/24/20 | 3/16/20 | 3/24/20 |
| Oklahoma | 3/17/20 | 3/29/20 | 3/16/20 | 3/29/20 | 3/16/20 | 3/29/20 |
| Oregon | 3/14/20 | 3/24/20 | 3/16/20 | 3/24/20 | 3/16/20 | 3/24/20 |
| Pennsylvania | 3/14/20 | 4/2/20 | 3/16/20 | 4/2/20 | 3/16/20 | 4/2/20 |
| Puerto_Rico | 3/14/20 | 3/16/20 | 3/16/20 | 3/16/20 | 3/16/20 | 3/16/20 |
| Rhode_Island | 3/21/20 | 3/29/20 | 3/16/20 | 3/29/20 | 3/16/20 | 3/29/20 |
| South_Carolina | 3/14/20 | 3/29/20 | 3/16/20 | 4/8/20 | 3/16/20 | 3/29/20 |
| Tennessee | 3/20/20 | 4/1/20 | 3/16/20 | 4/1/20 | 3/16/20 | 4/1/20 |
| Texas | 3/21/20 | 4/2/20 | 3/16/20 | 4/2/20 | 3/16/20 | 4/2/20 |
| Utah | 3/14/20 | 3/30/20 | 3/16/20 | 3/30/20 | 3/16/20 | 3/30/20 |
| Vermont | 3/18/20 | 3/26/20 | 3/16/20 | 3/26/20 | 3/16/20 | 3/26/20 |
| Virginia | 3/14/20 | 3/30/20 | 3/16/20 | 3/30/20 | 3/16/20 | 3/30/20 |
| Washington | 3/17/20 | 3/23/20 | 3/16/20 | 3/23/20 | 3/16/20 | 3/23/20 |
| West_Virginia | 3/14/20 | 3/25/20 | 3/16/20 | 3/25/20 | 3/16/20 | 3/25/20 |
| Wisconsin | 3/18/20 | 3/25/20 | 3/16/20 | 3/25/20 | 3/16/20 | 3/25/20 |

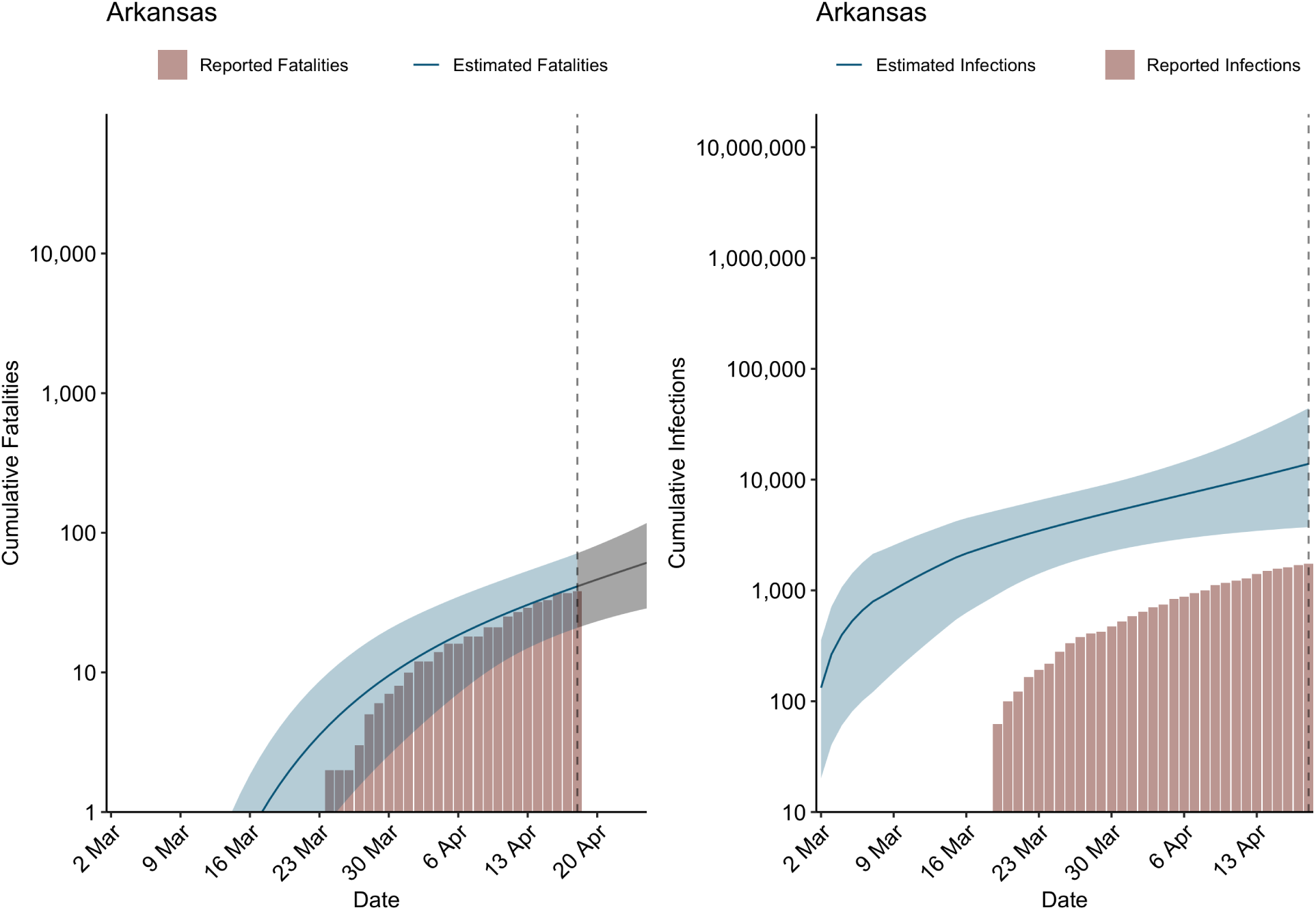
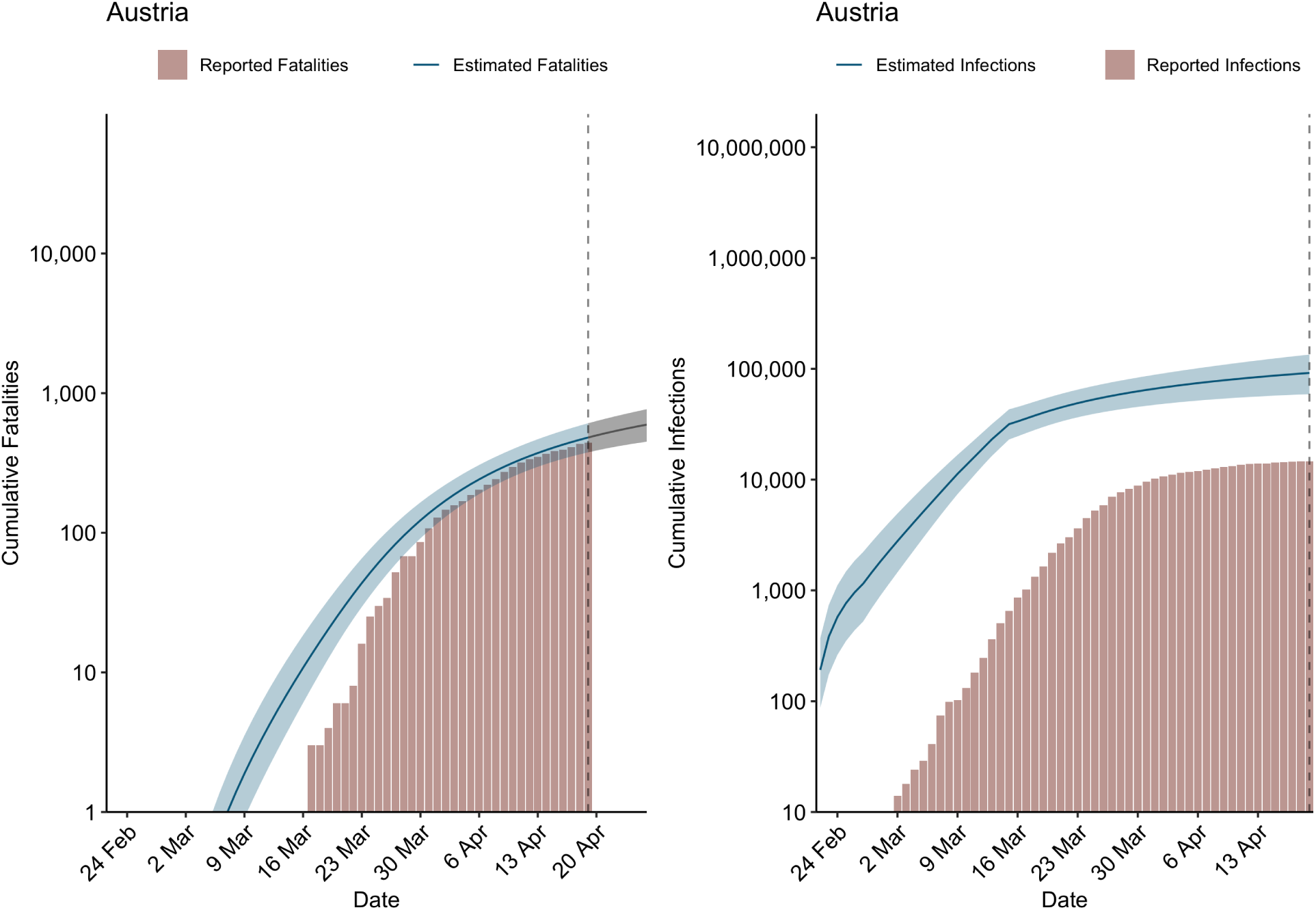
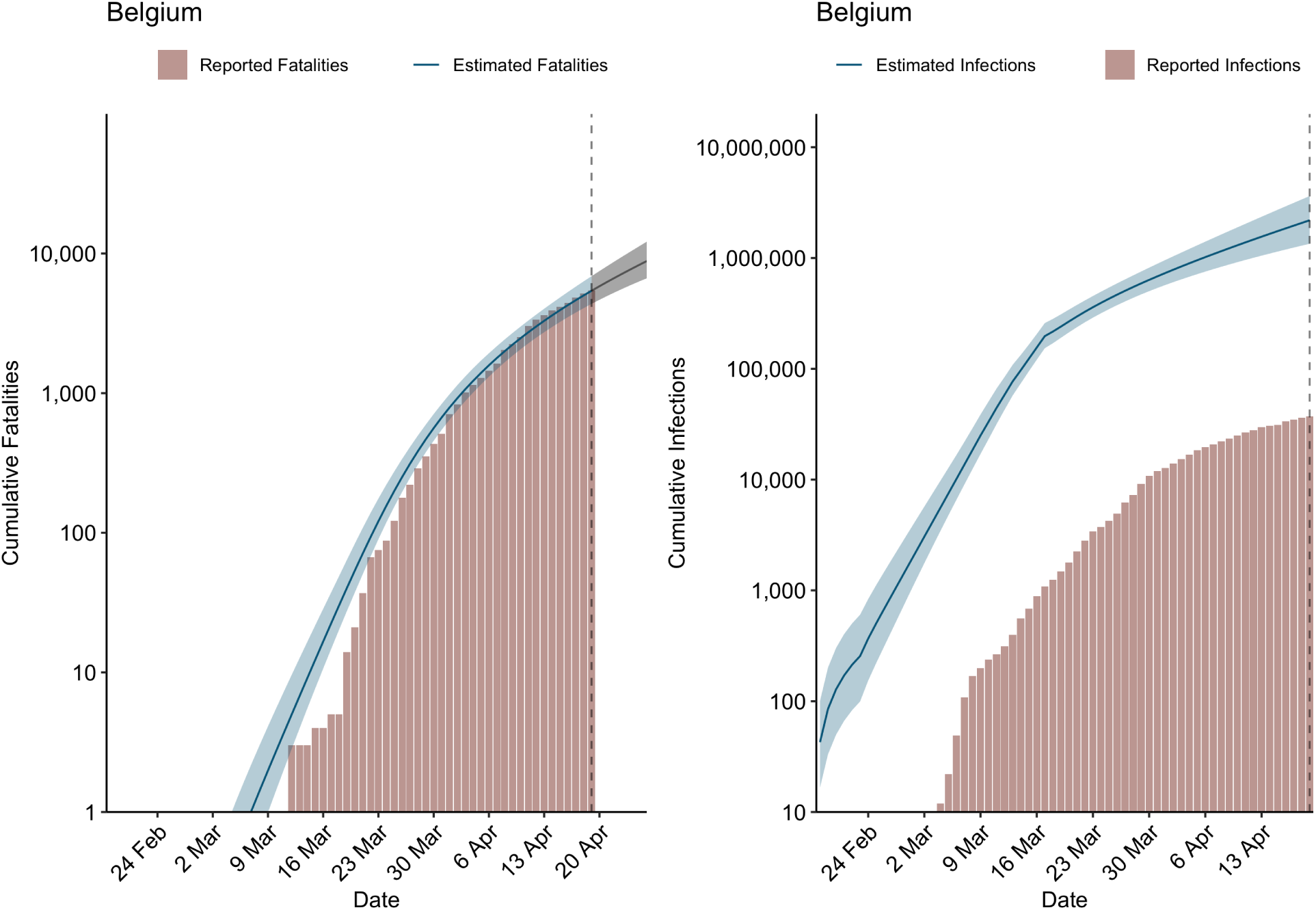
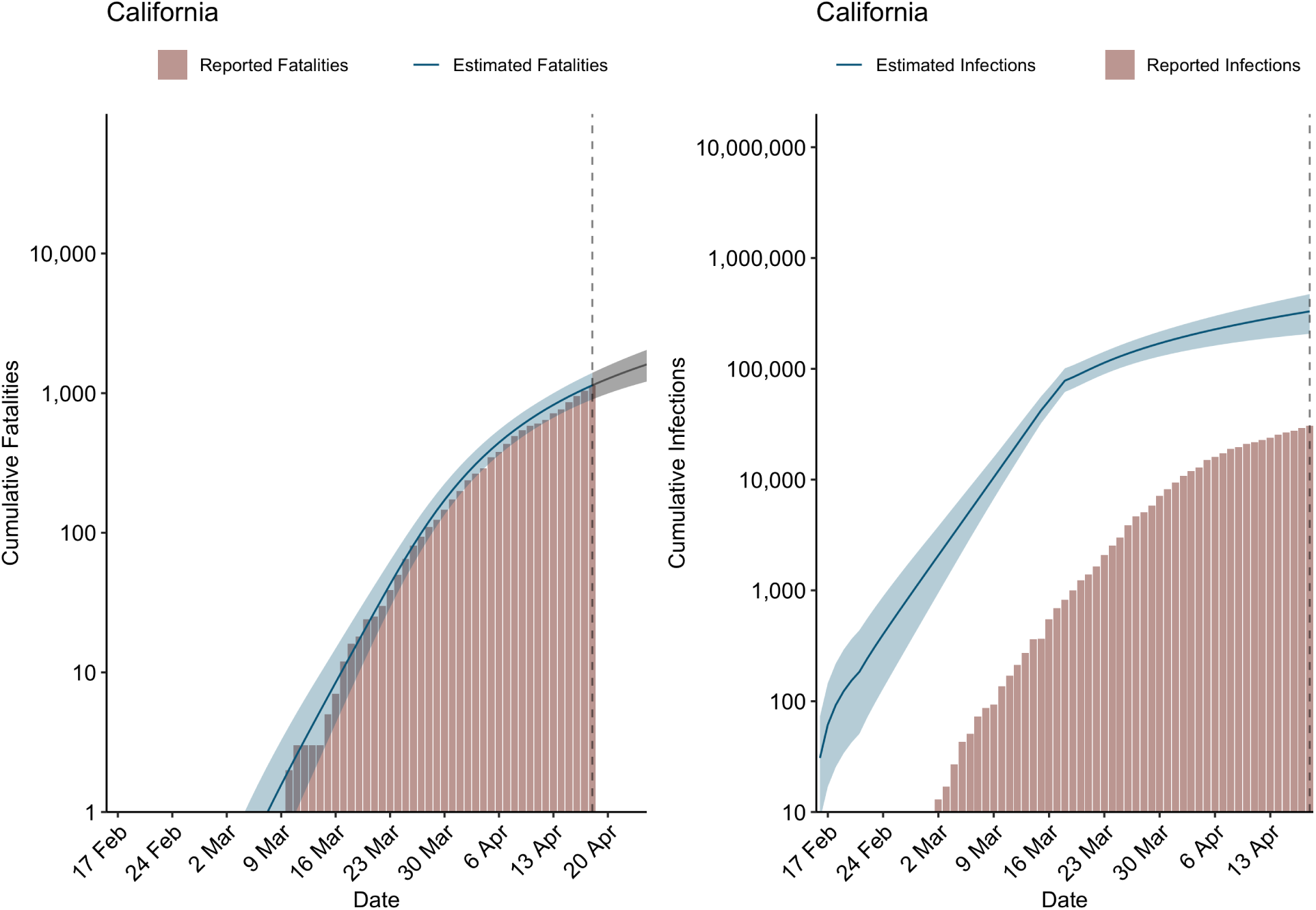
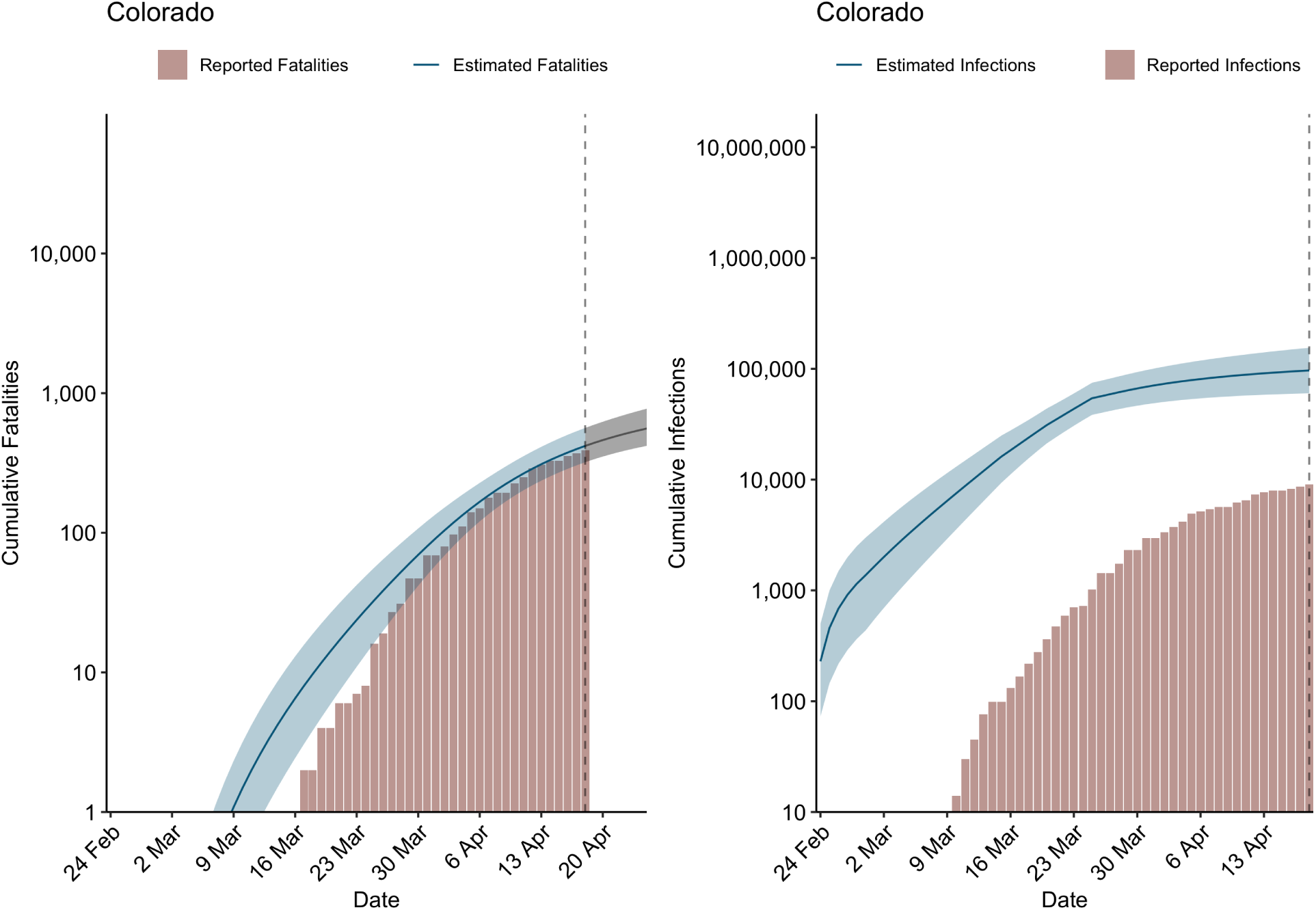
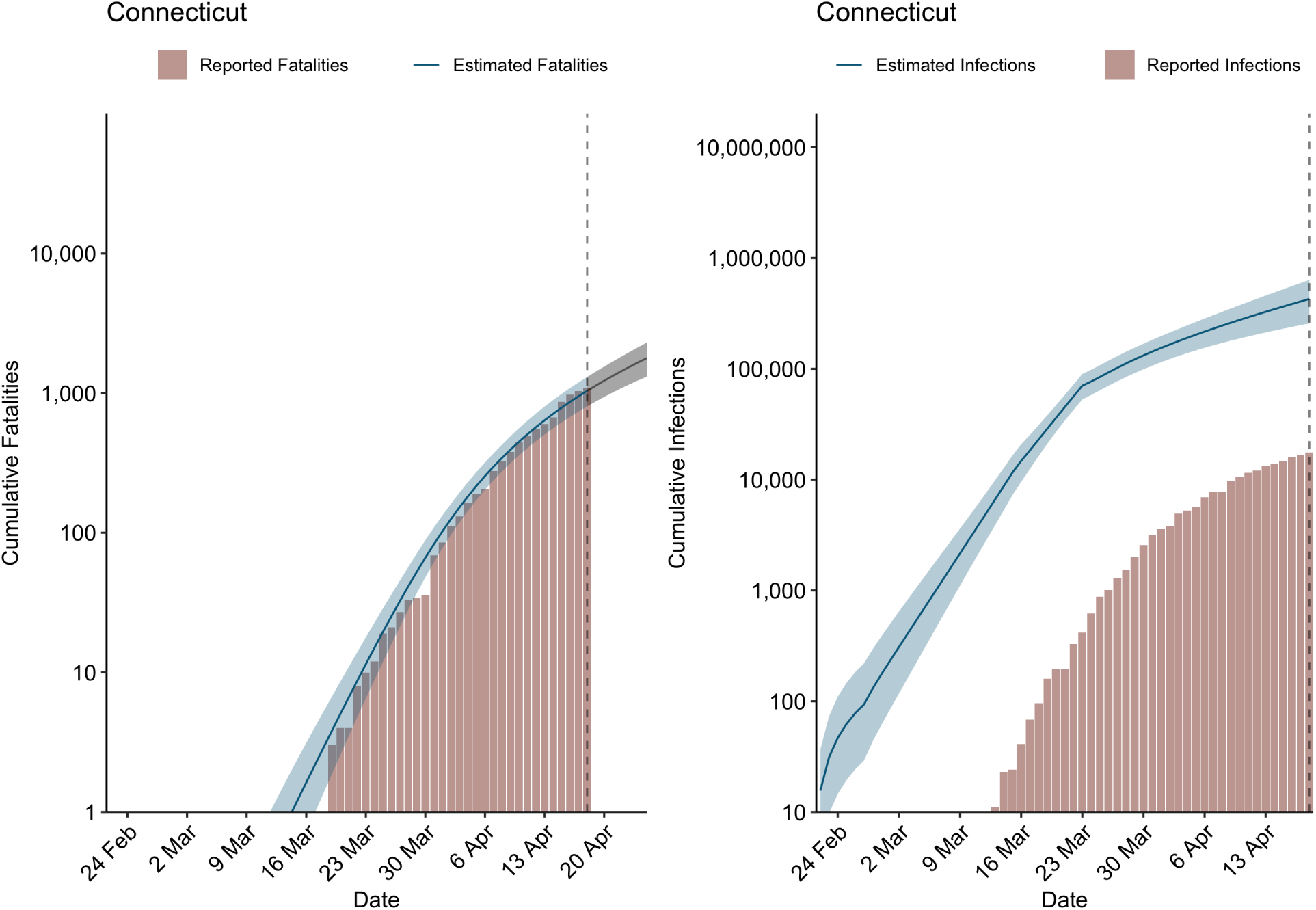
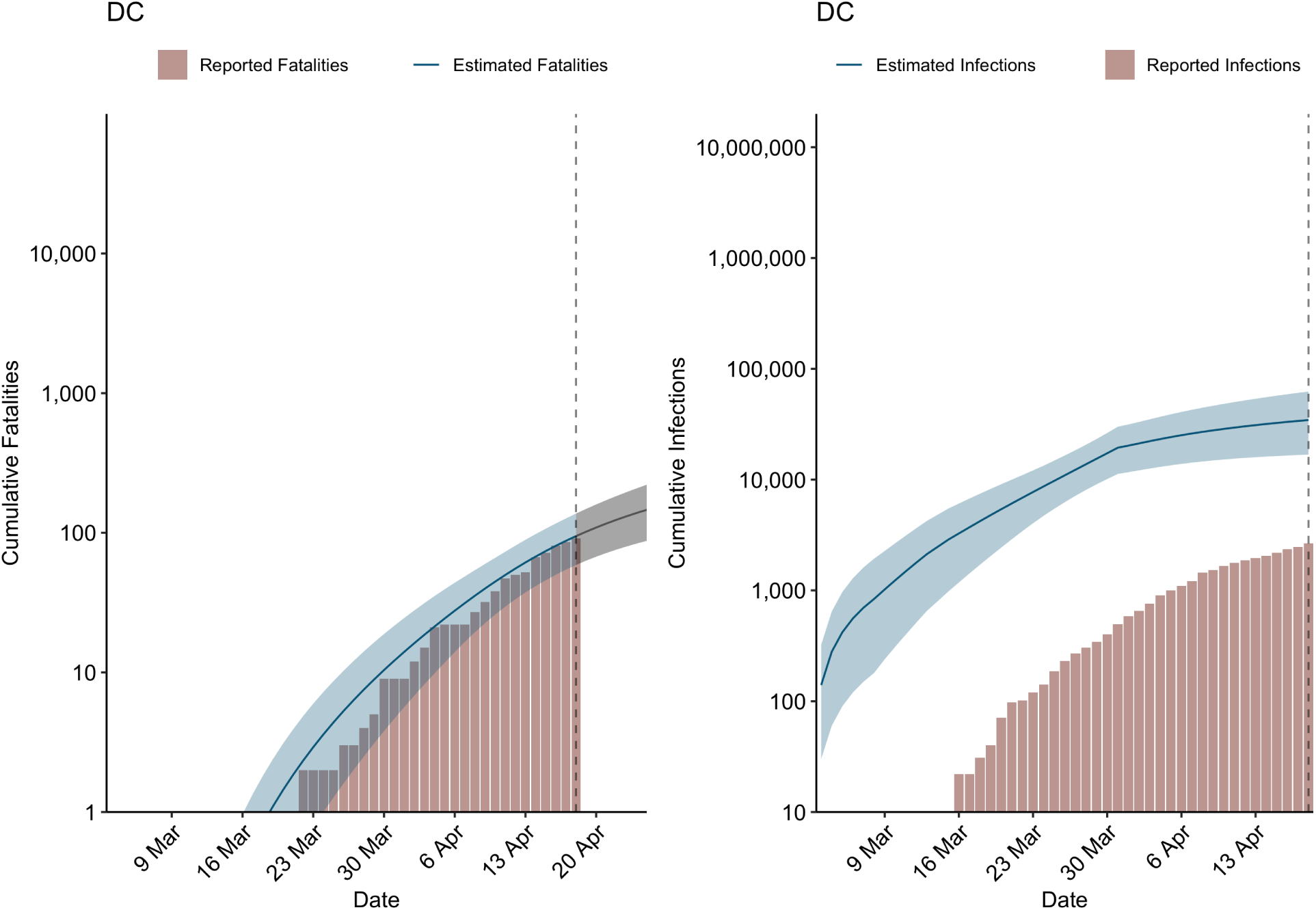
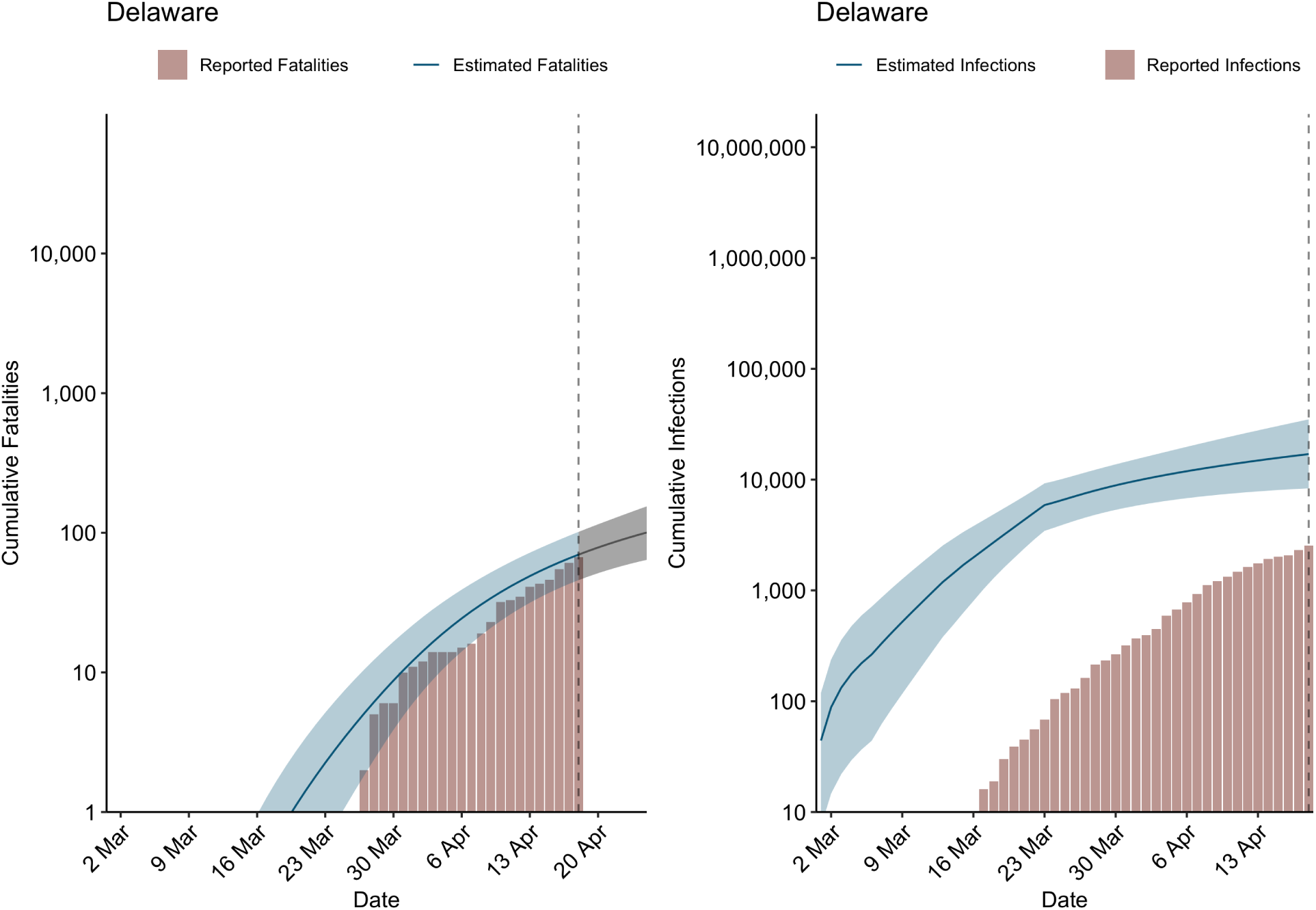
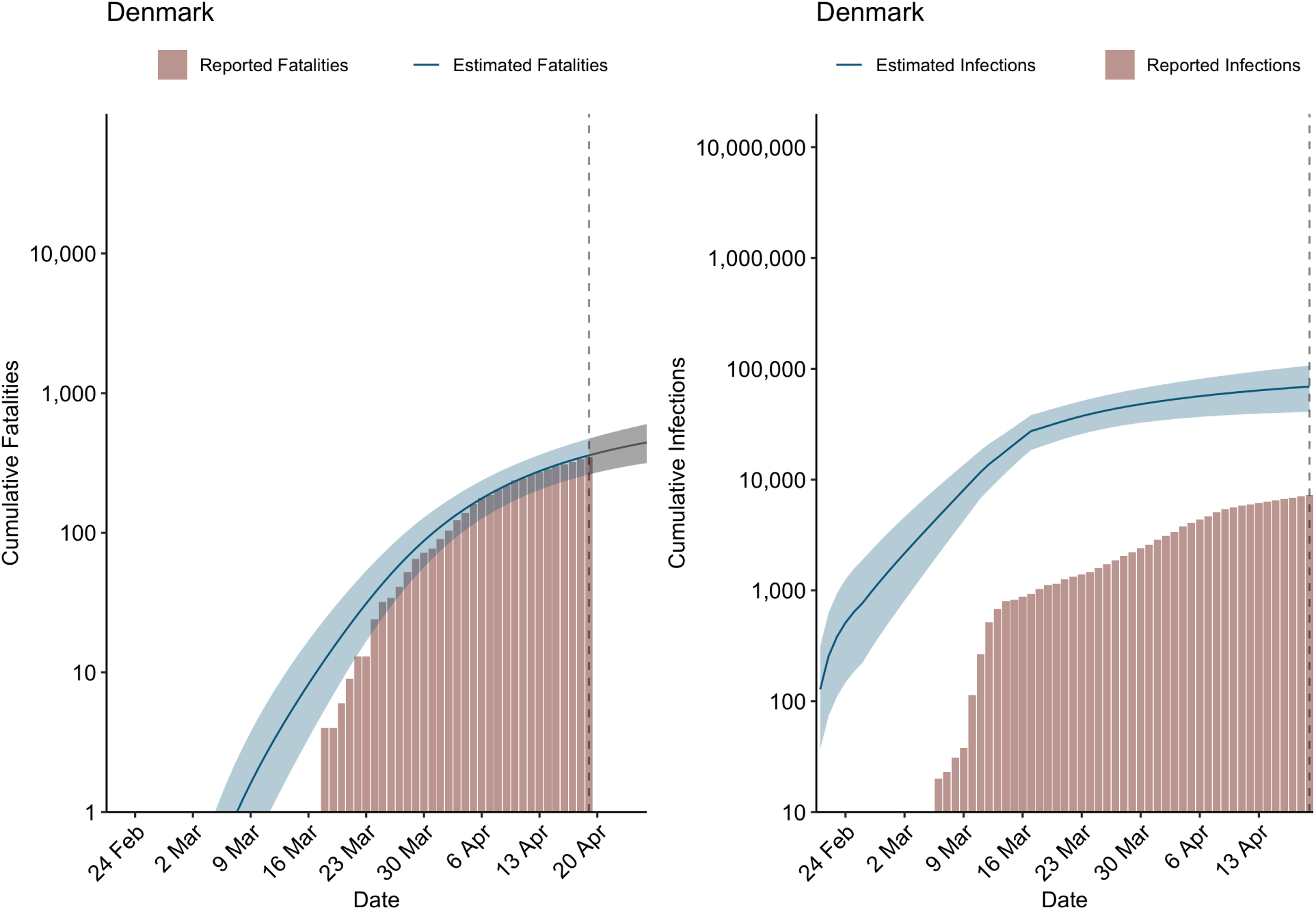
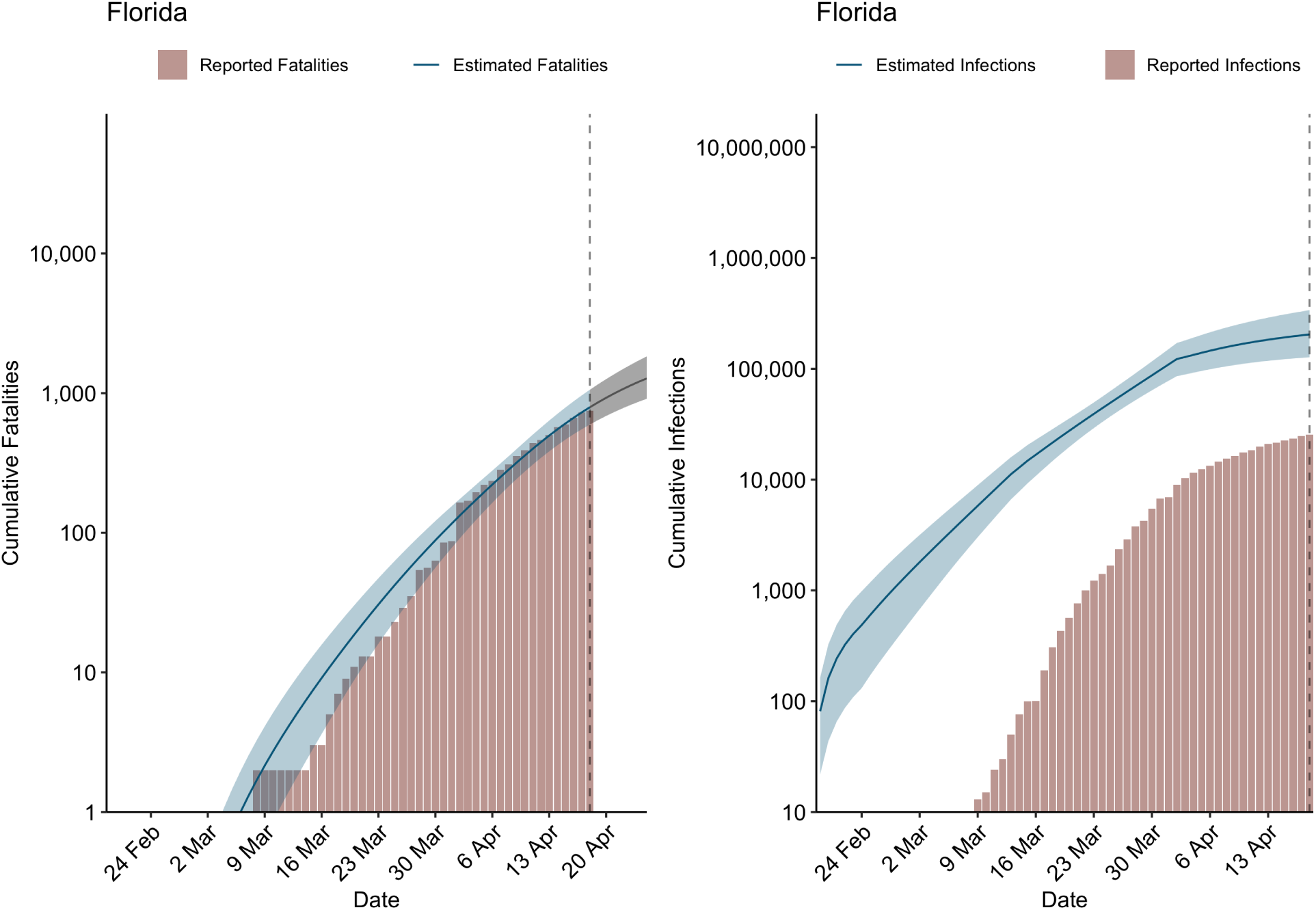
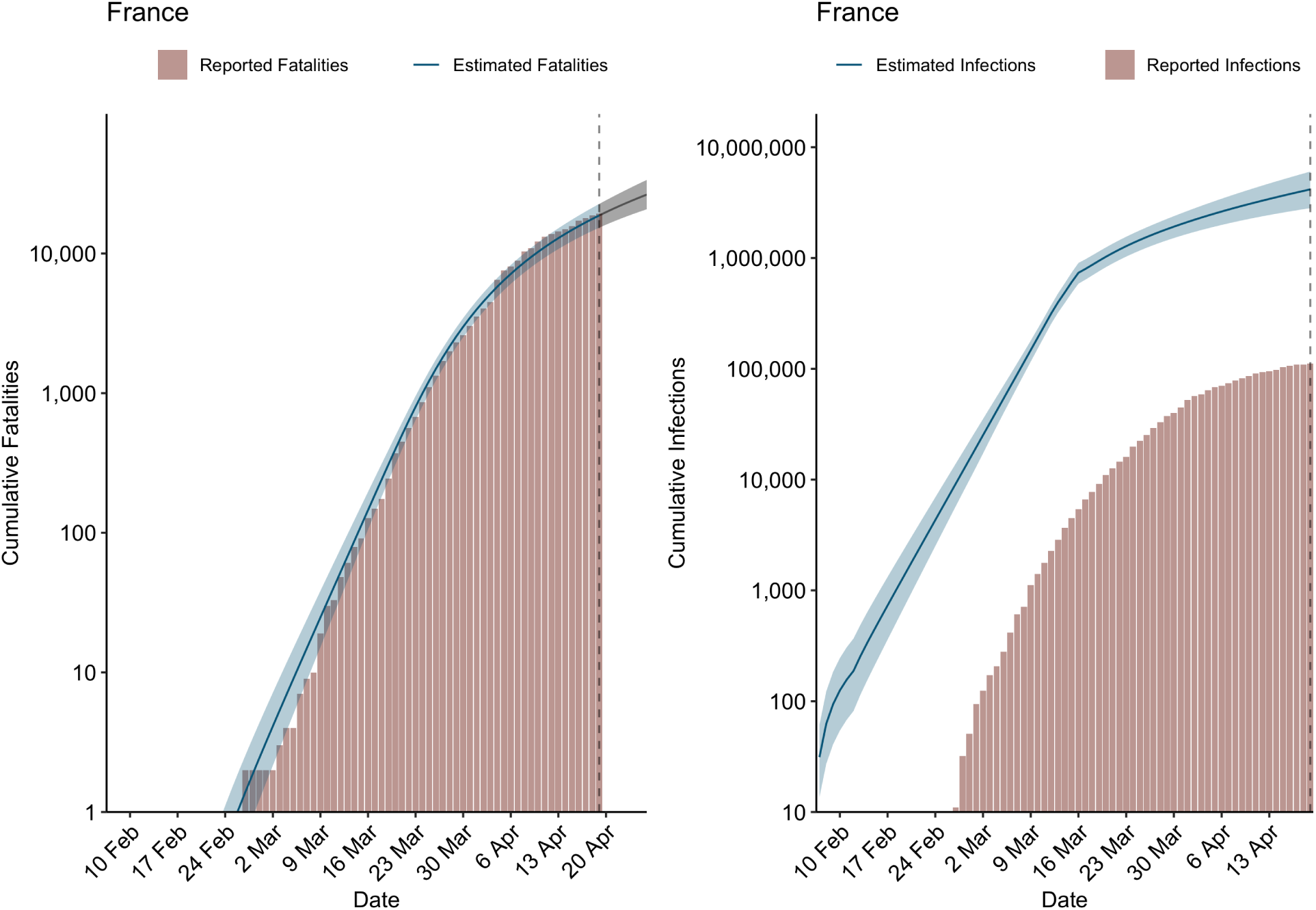
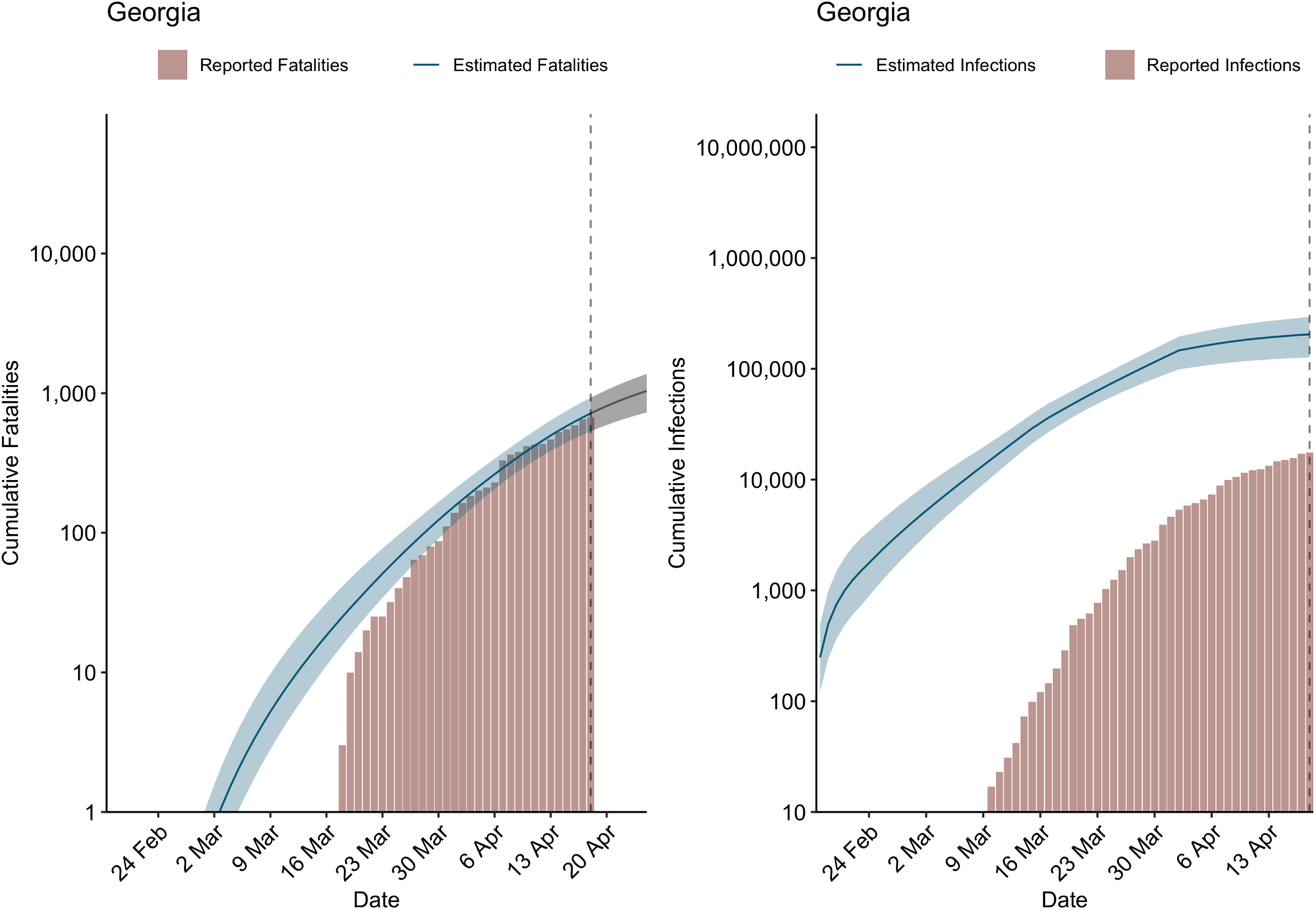
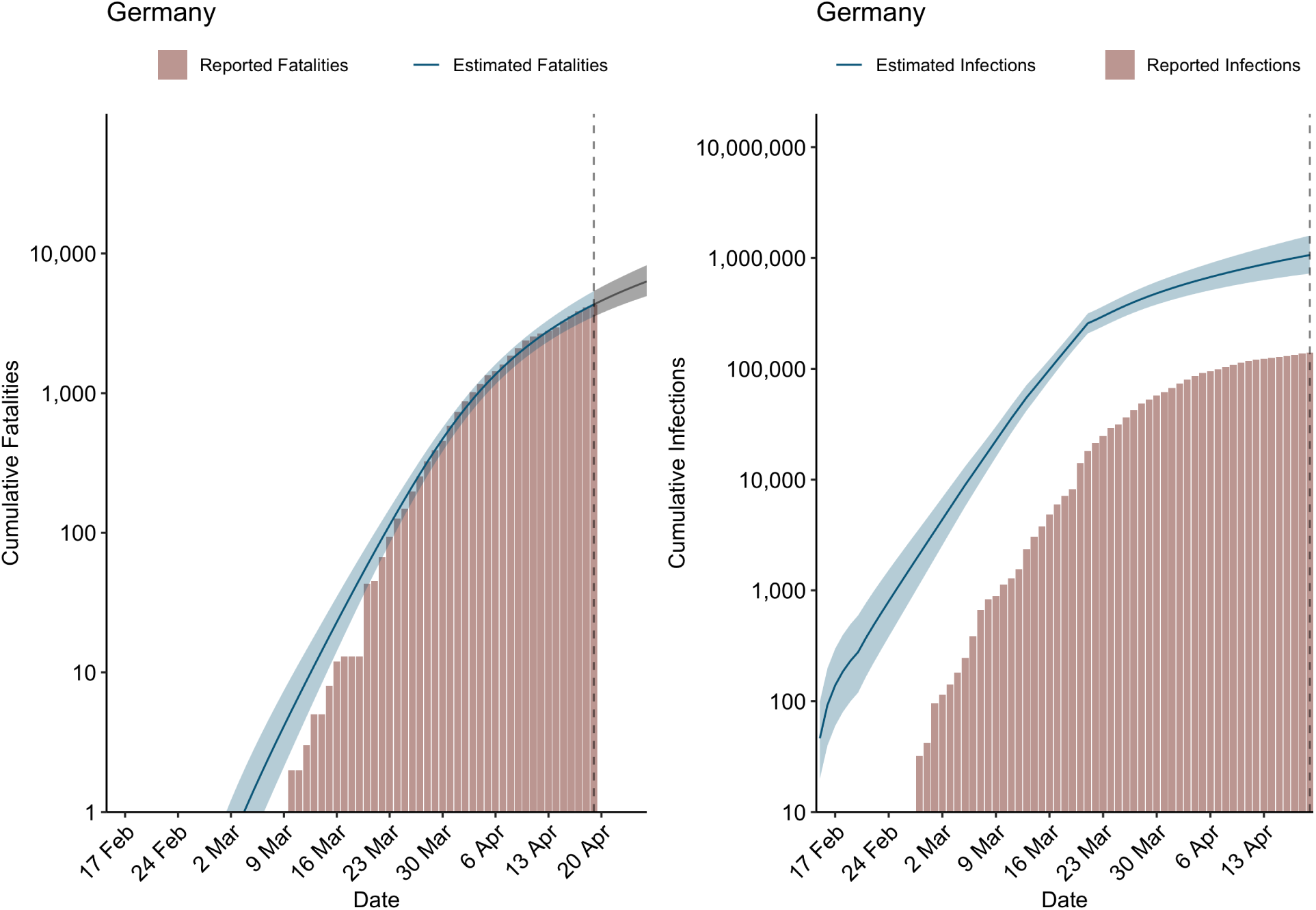
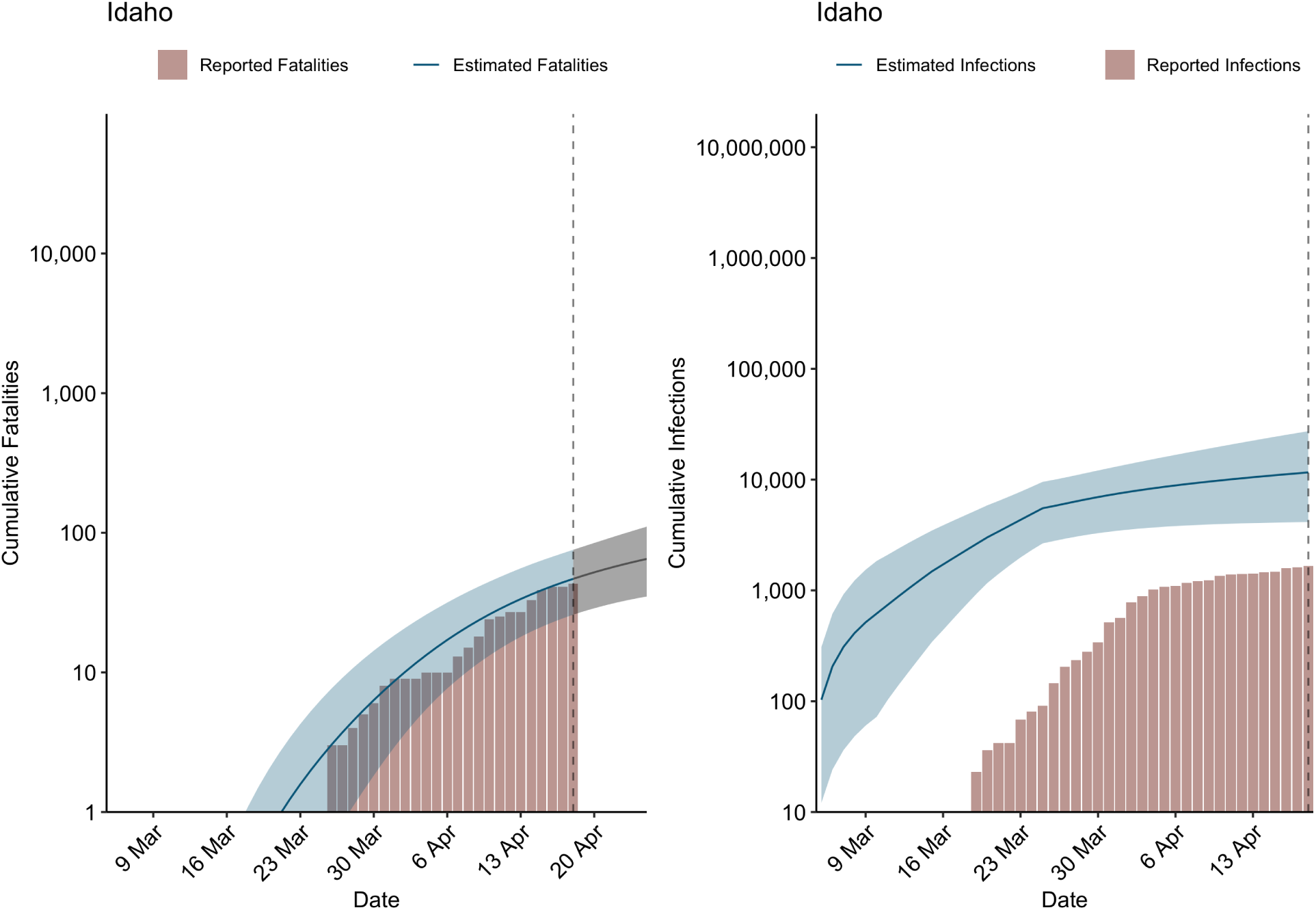
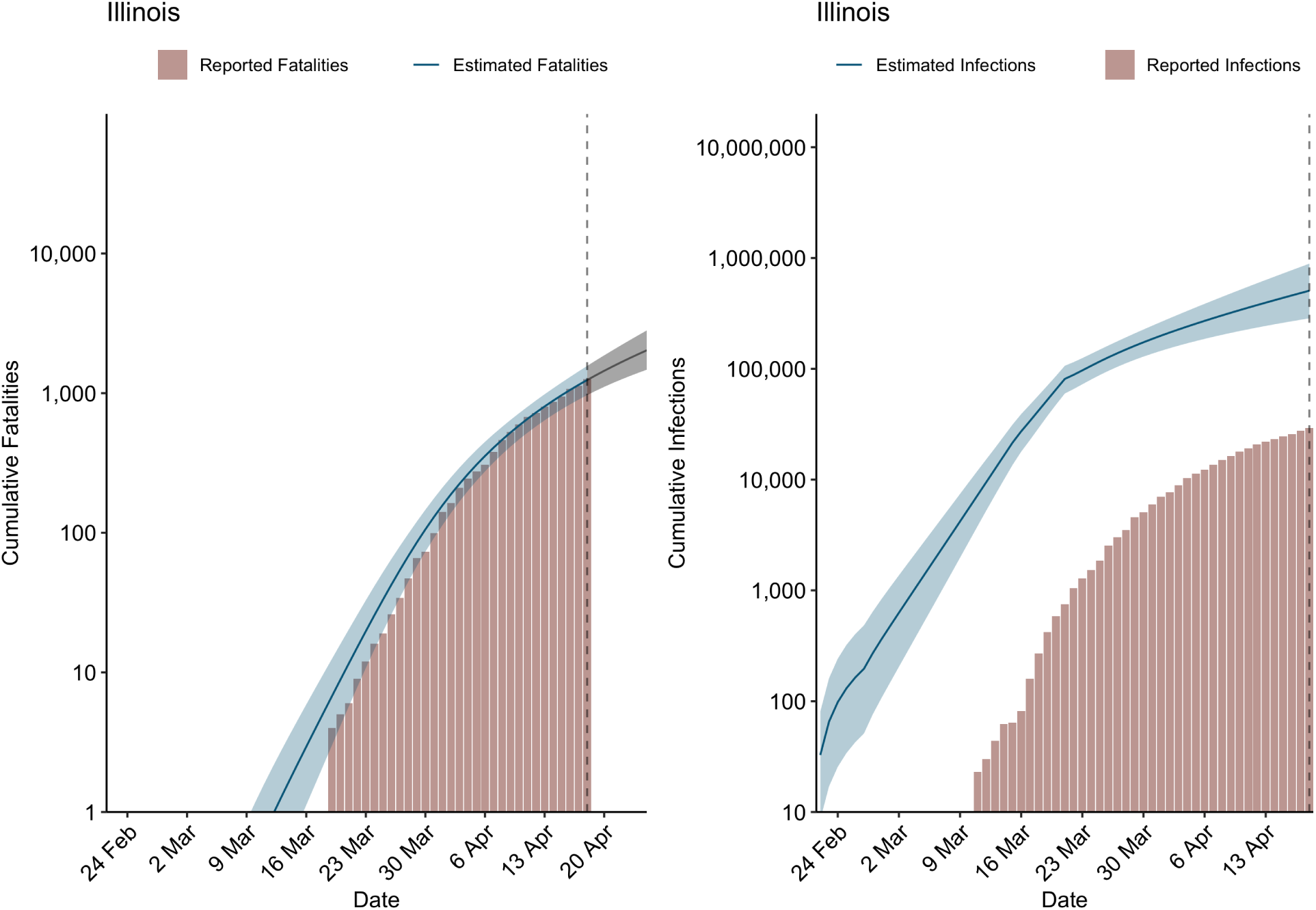
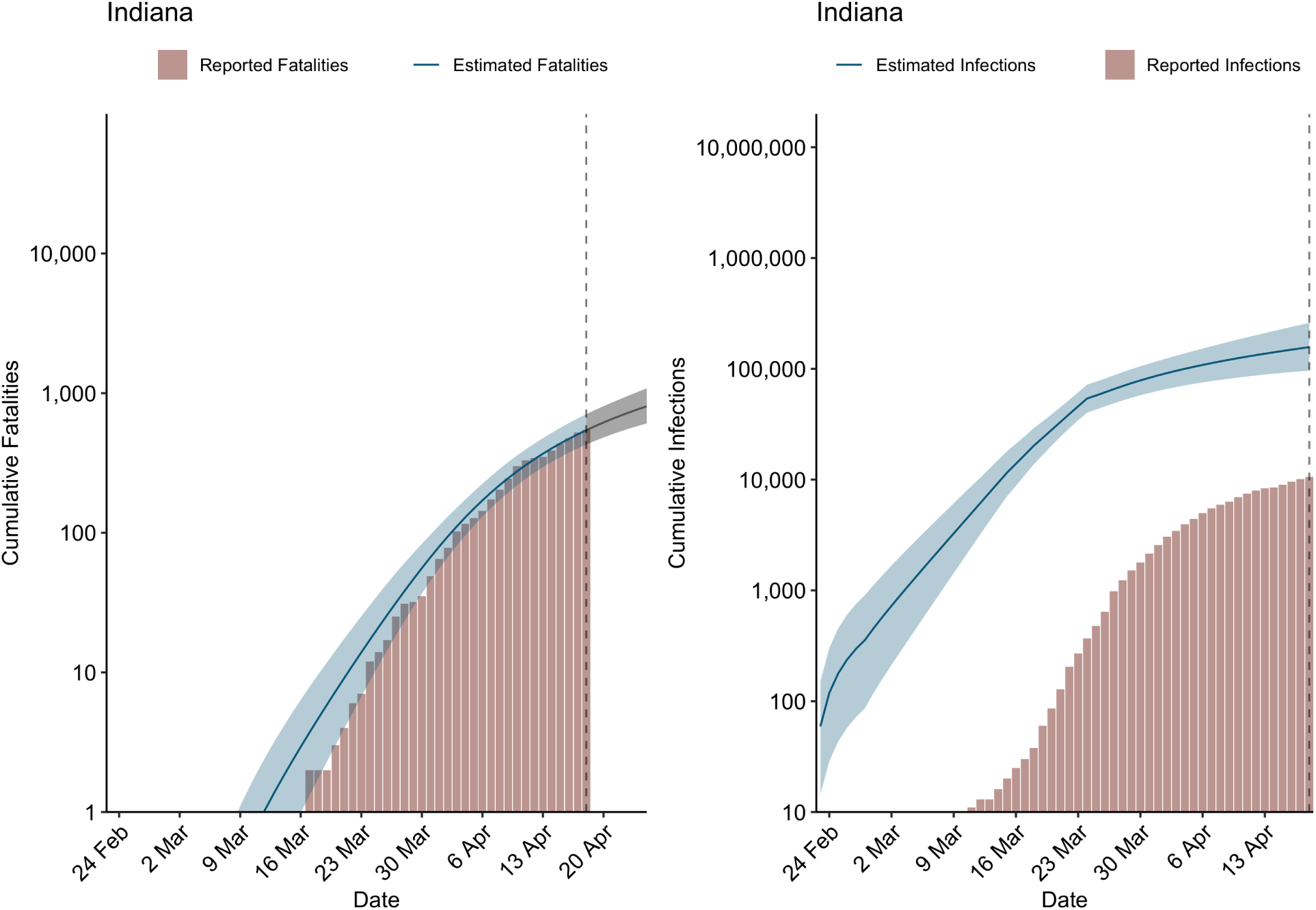
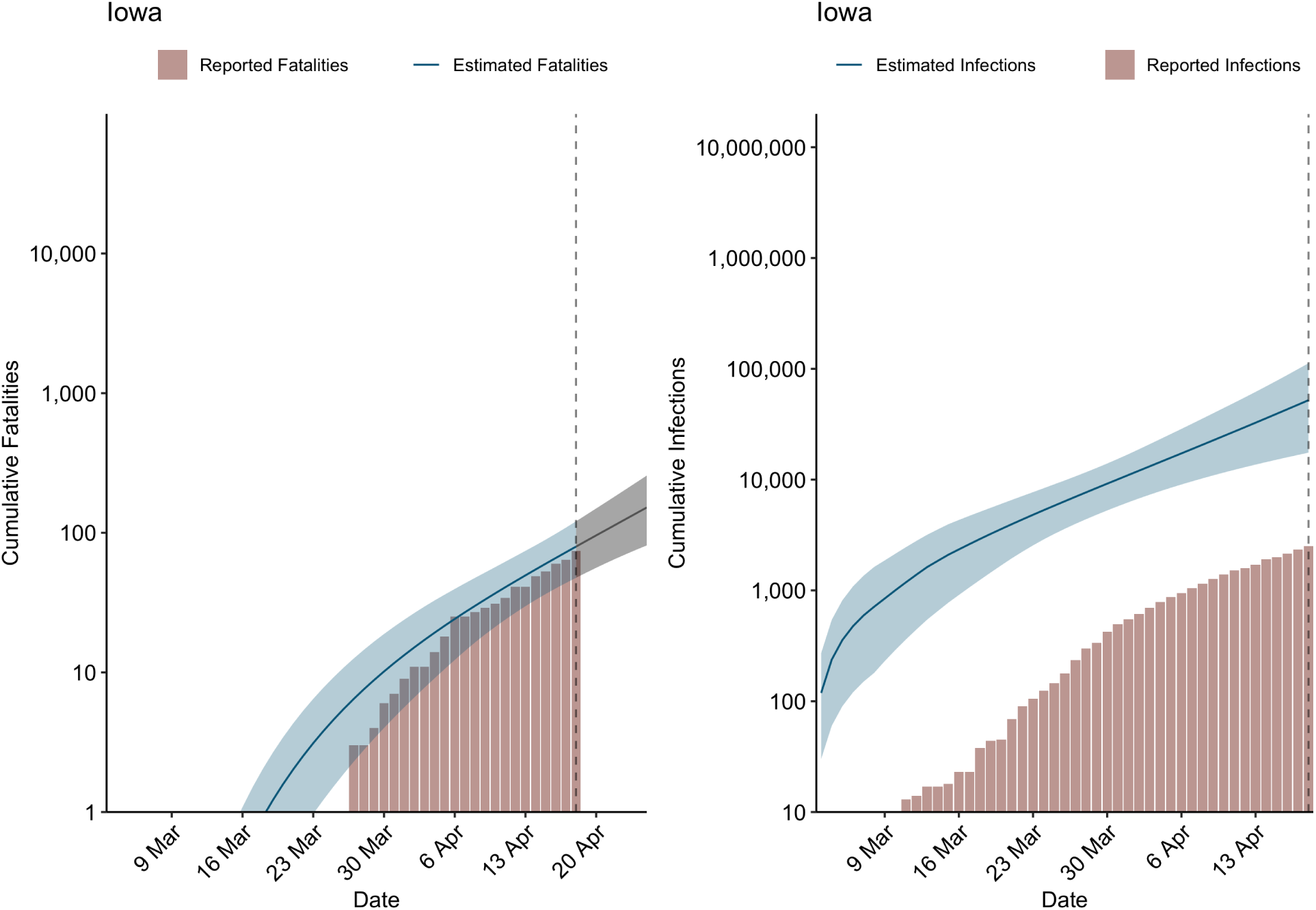
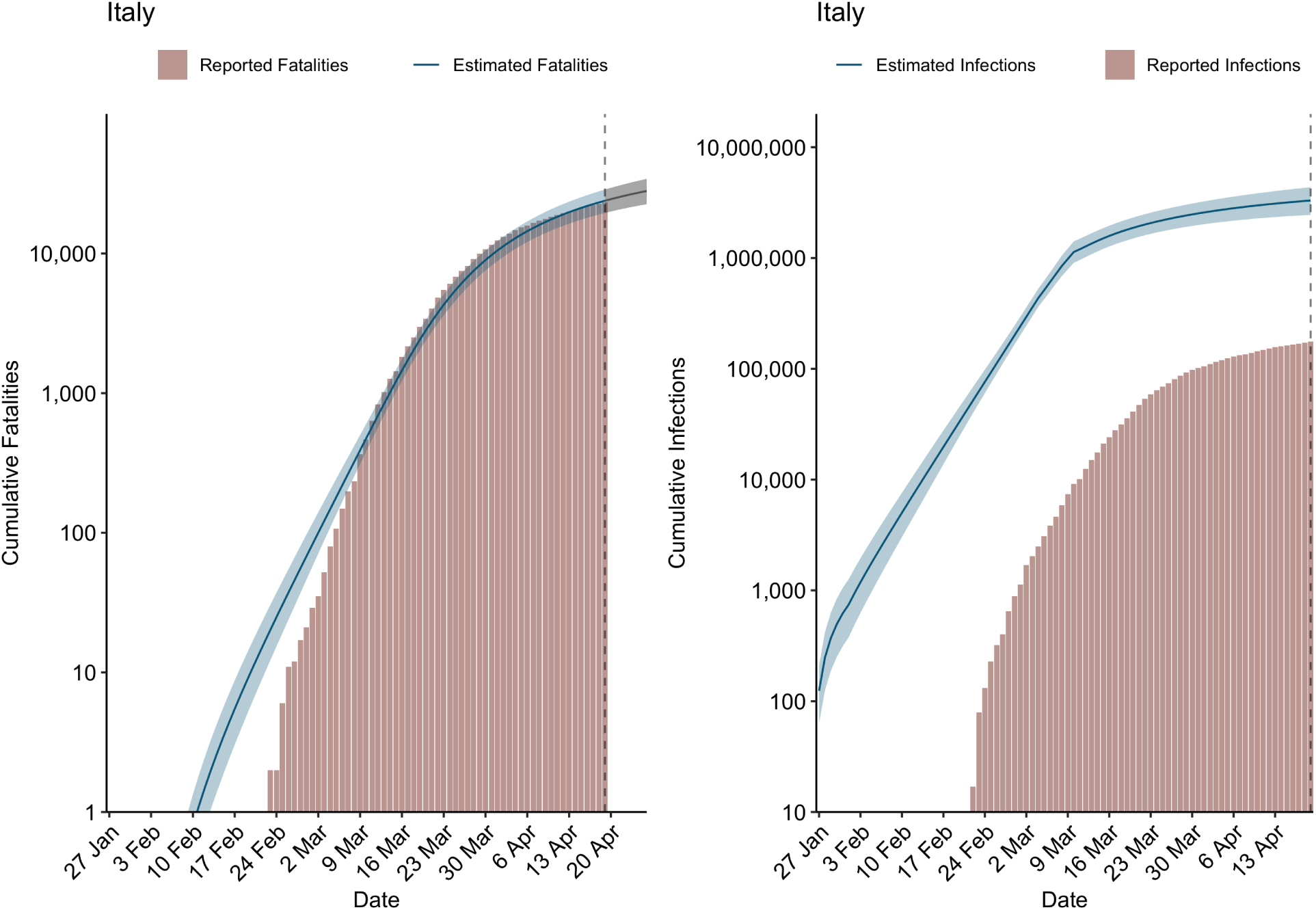
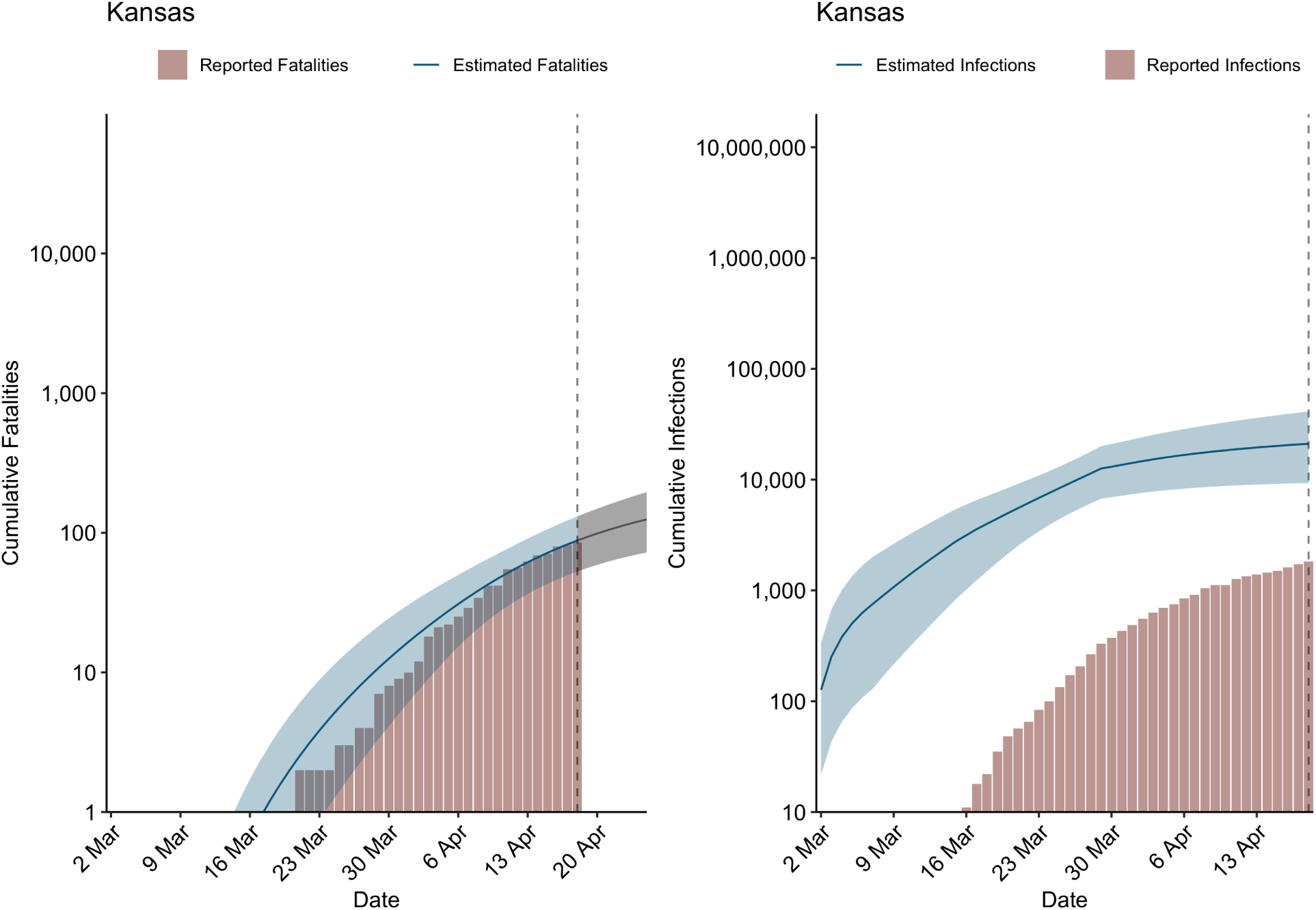
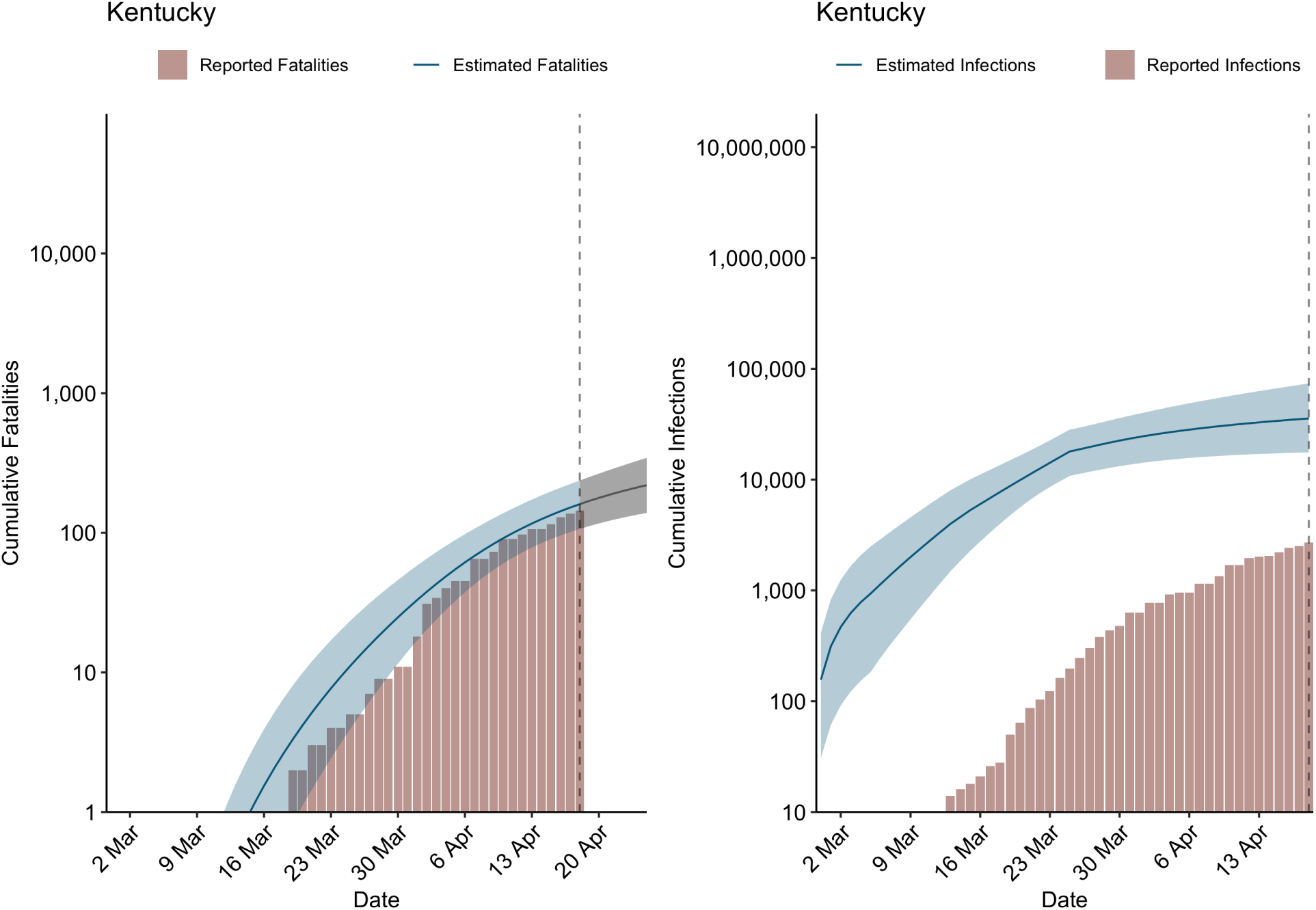
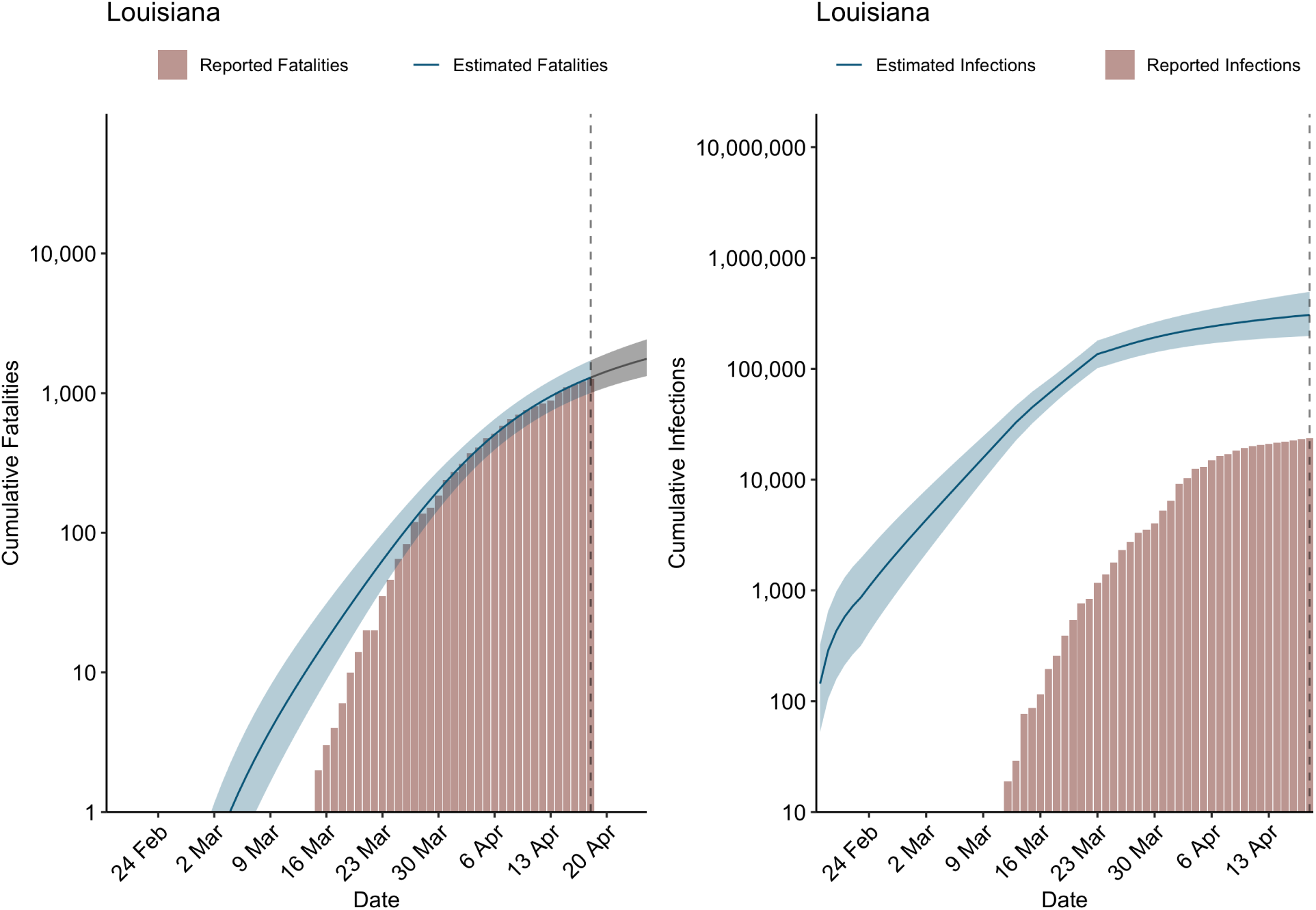
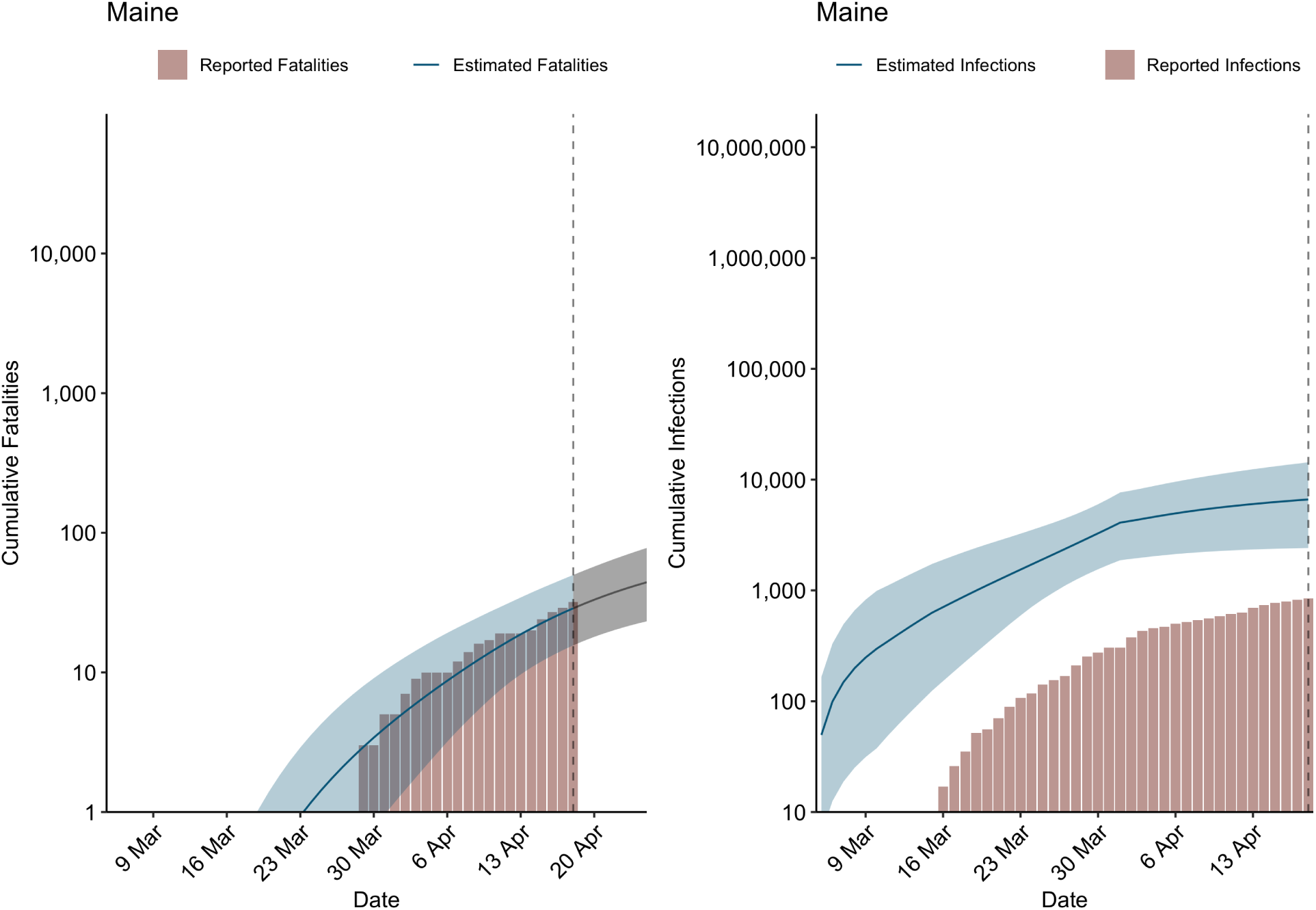
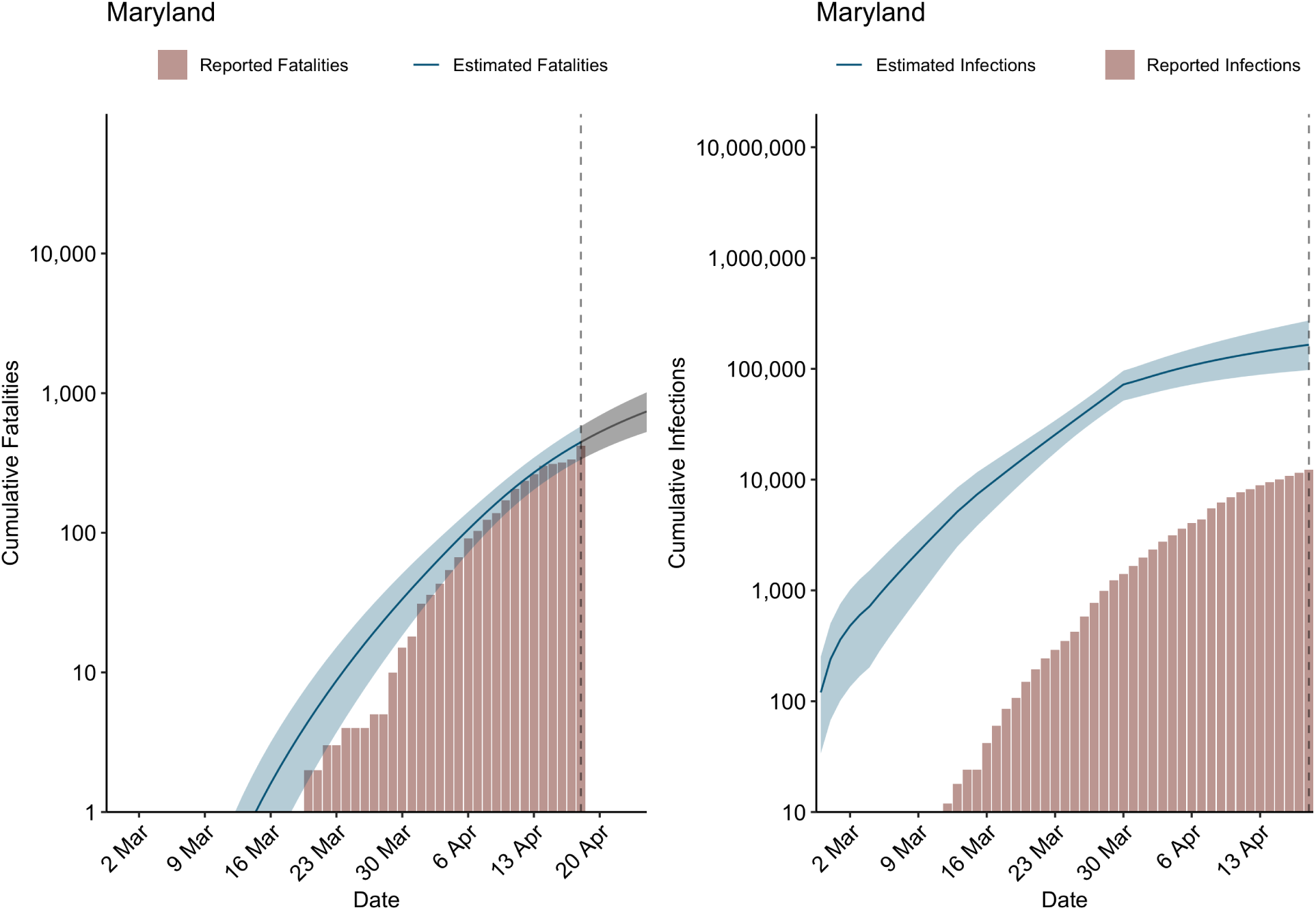
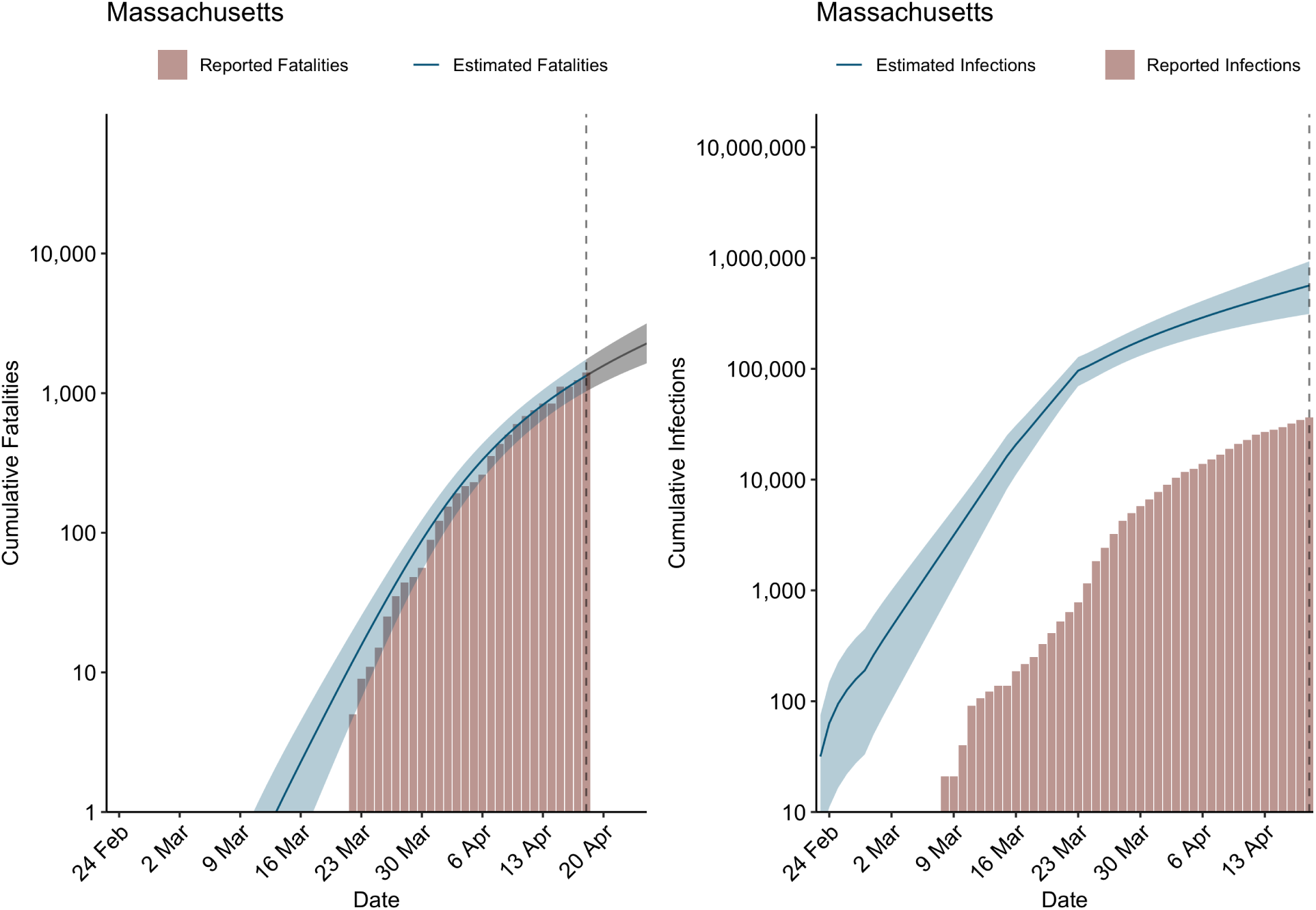
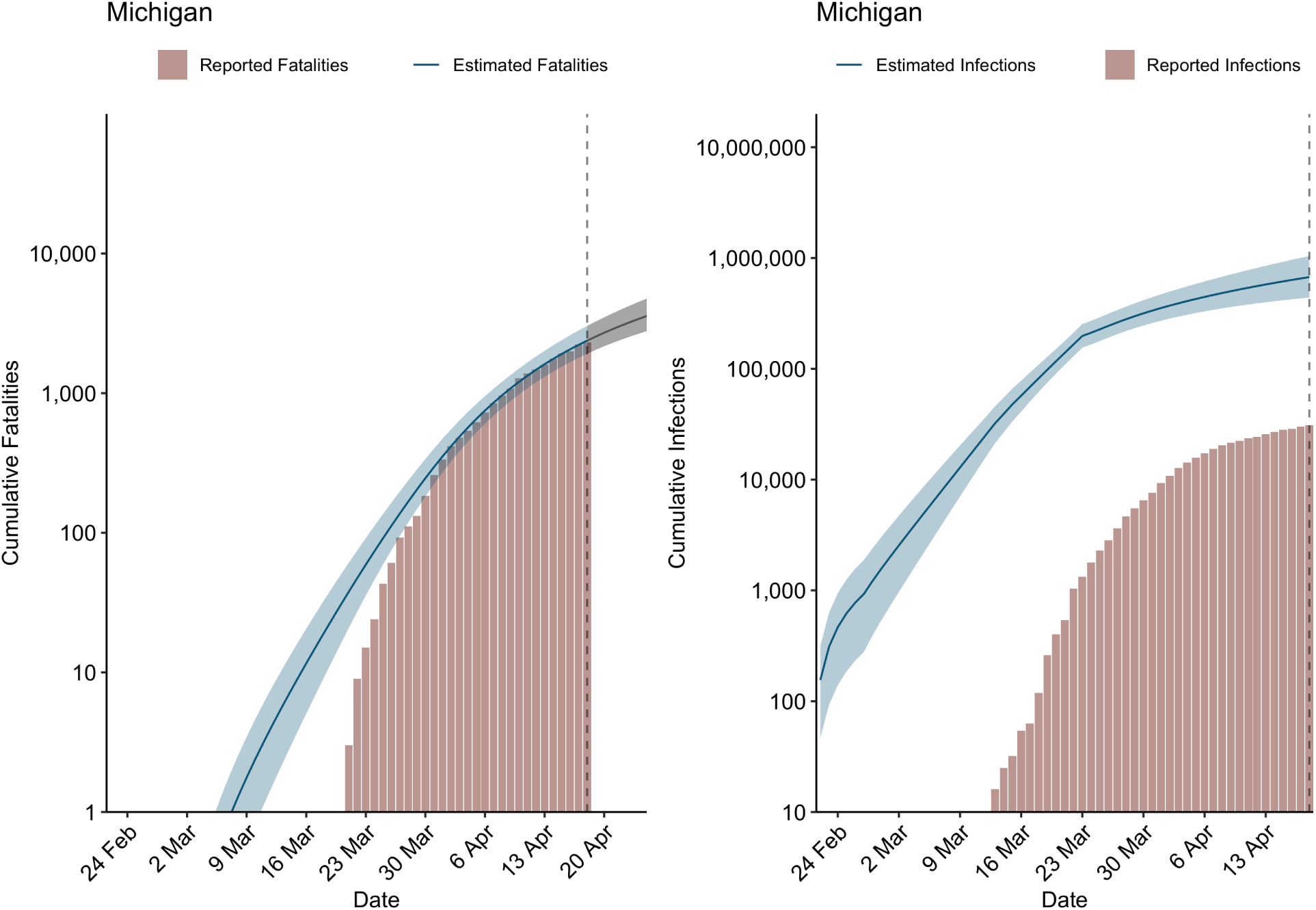
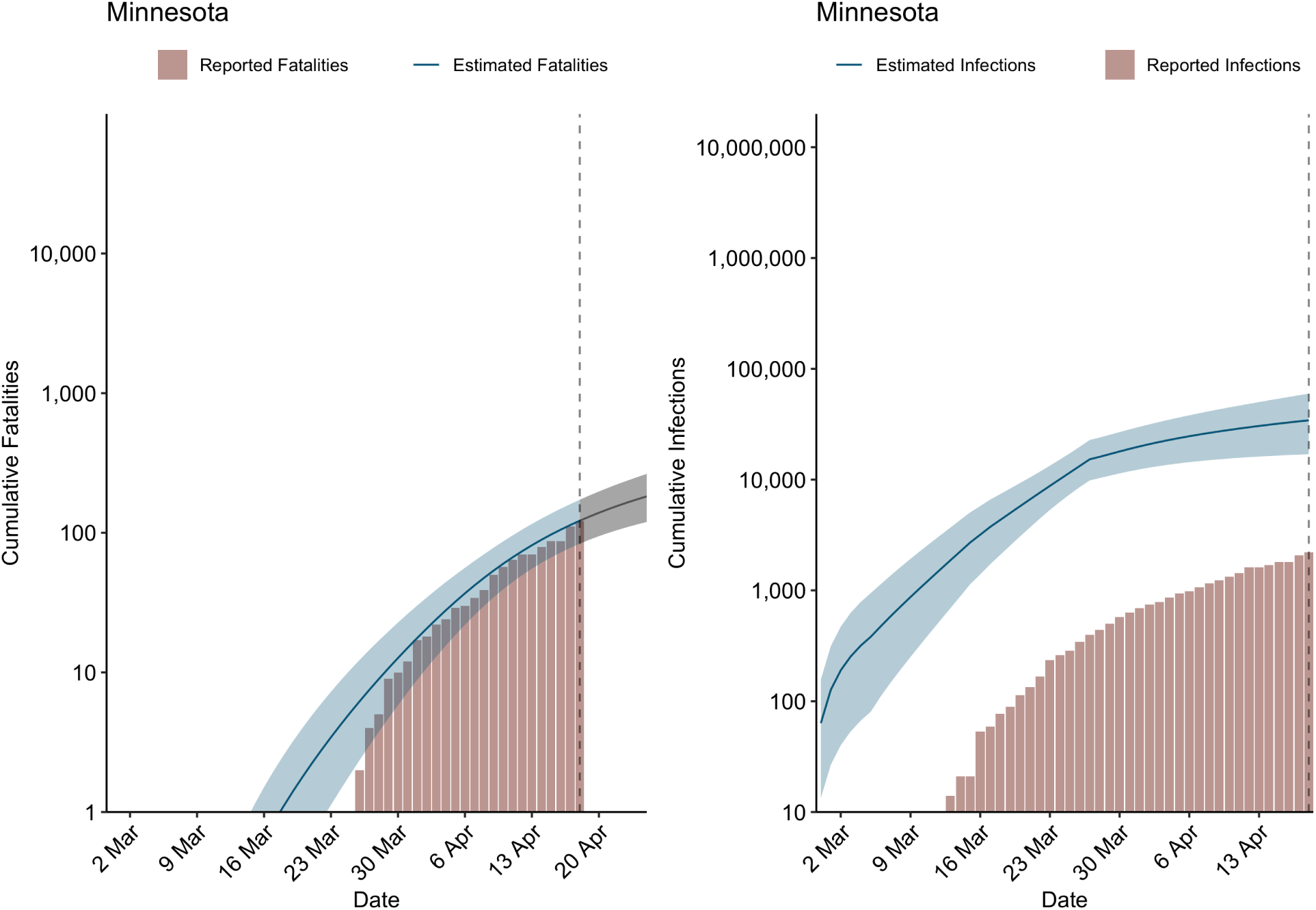
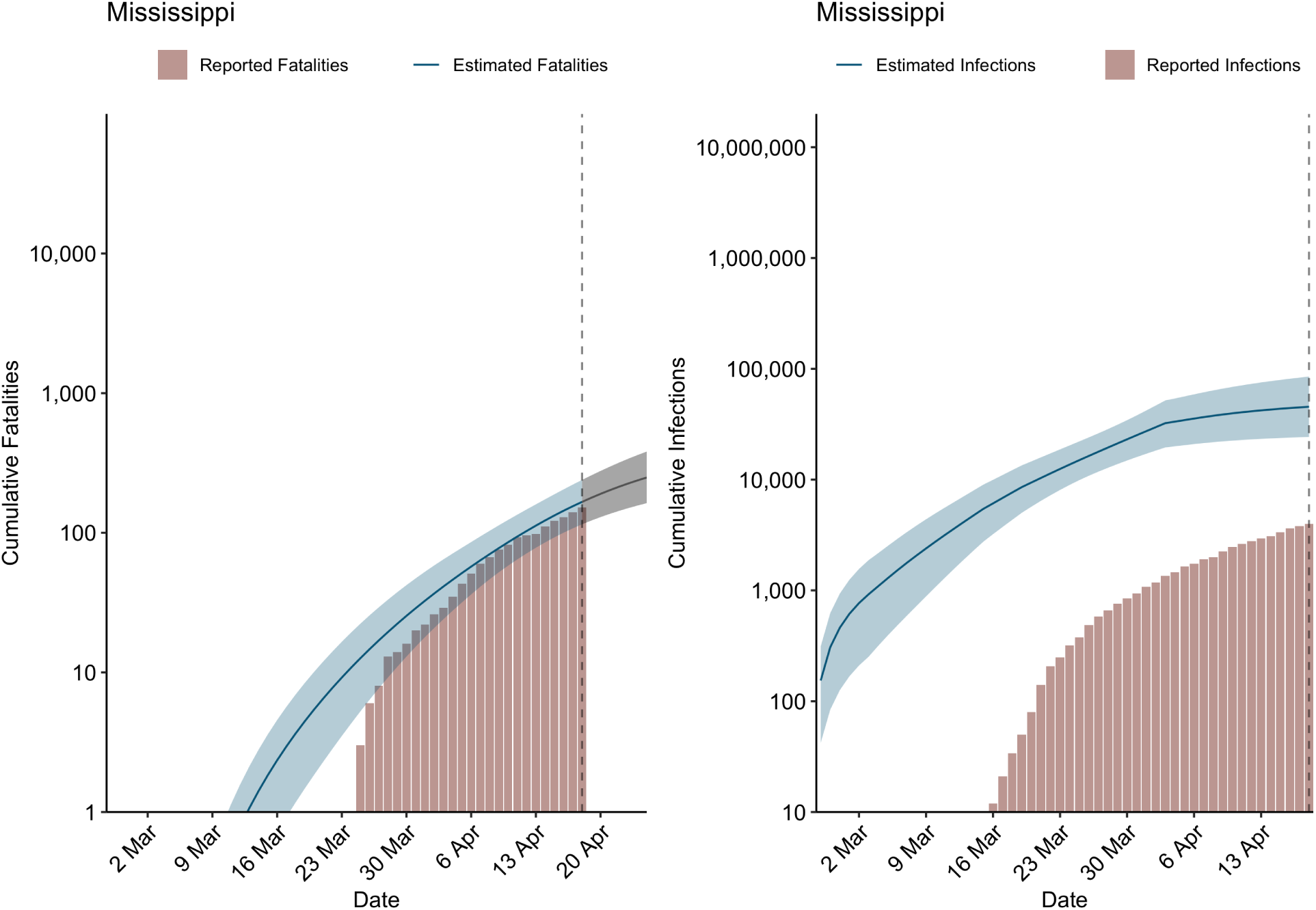
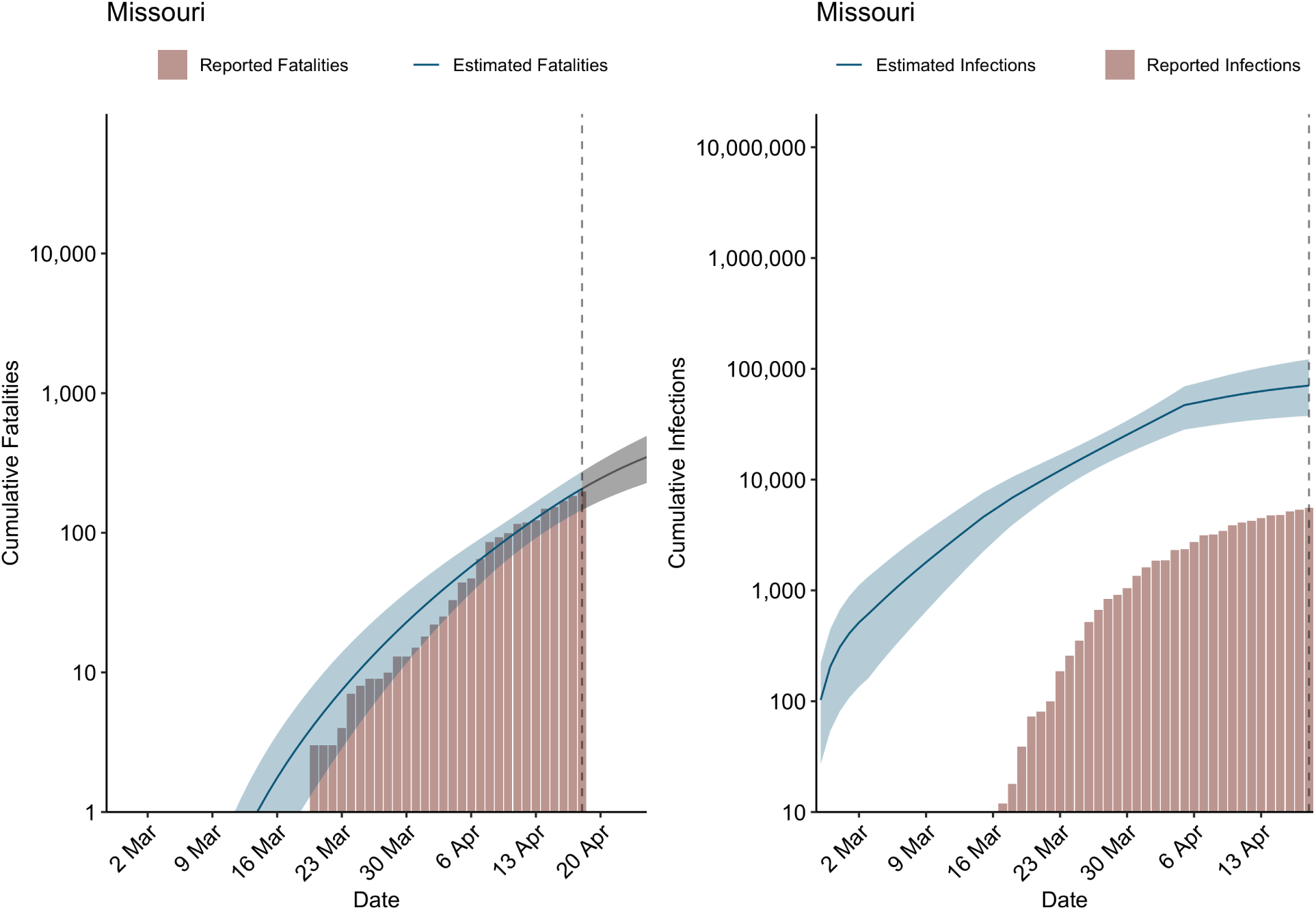
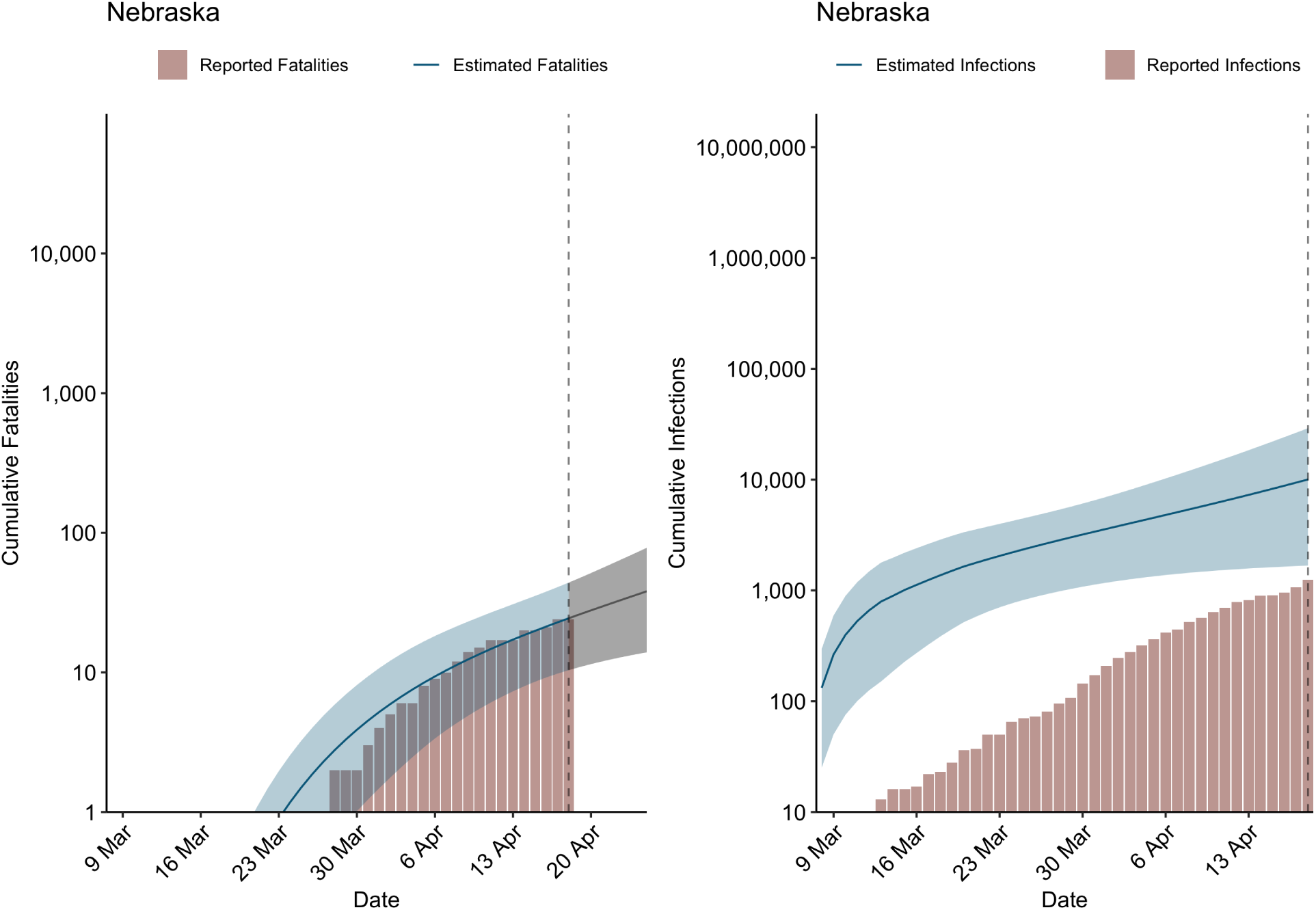
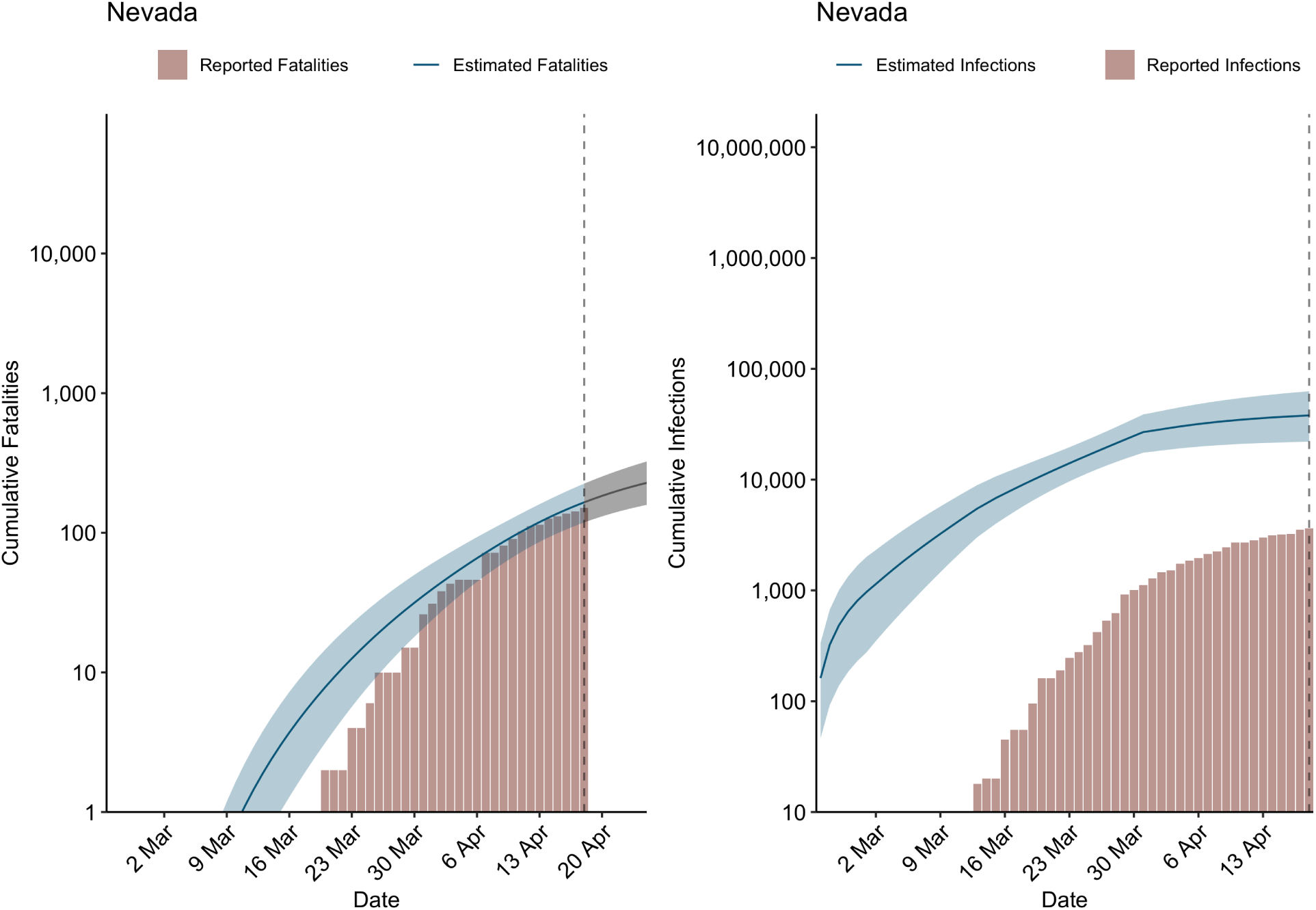
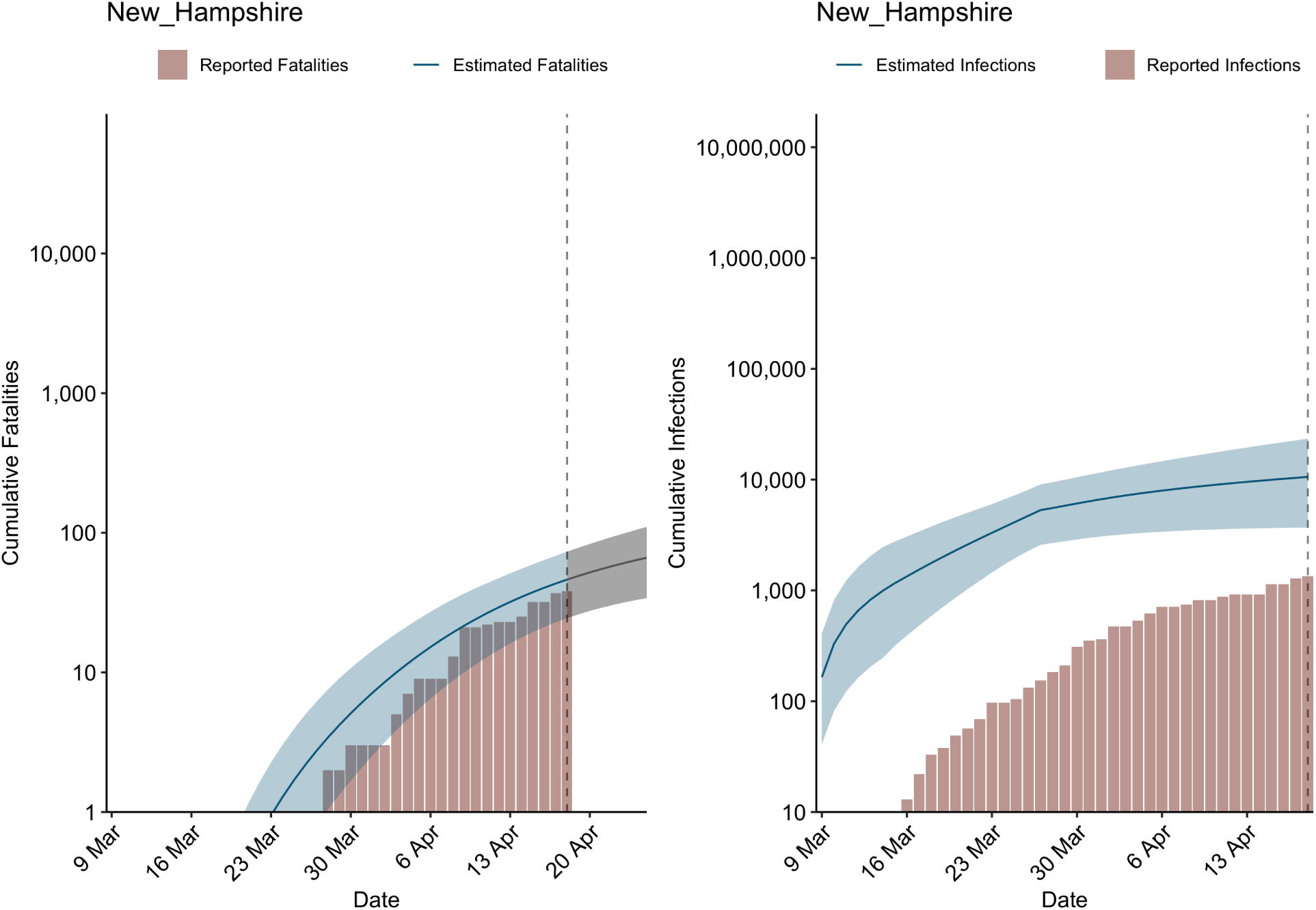
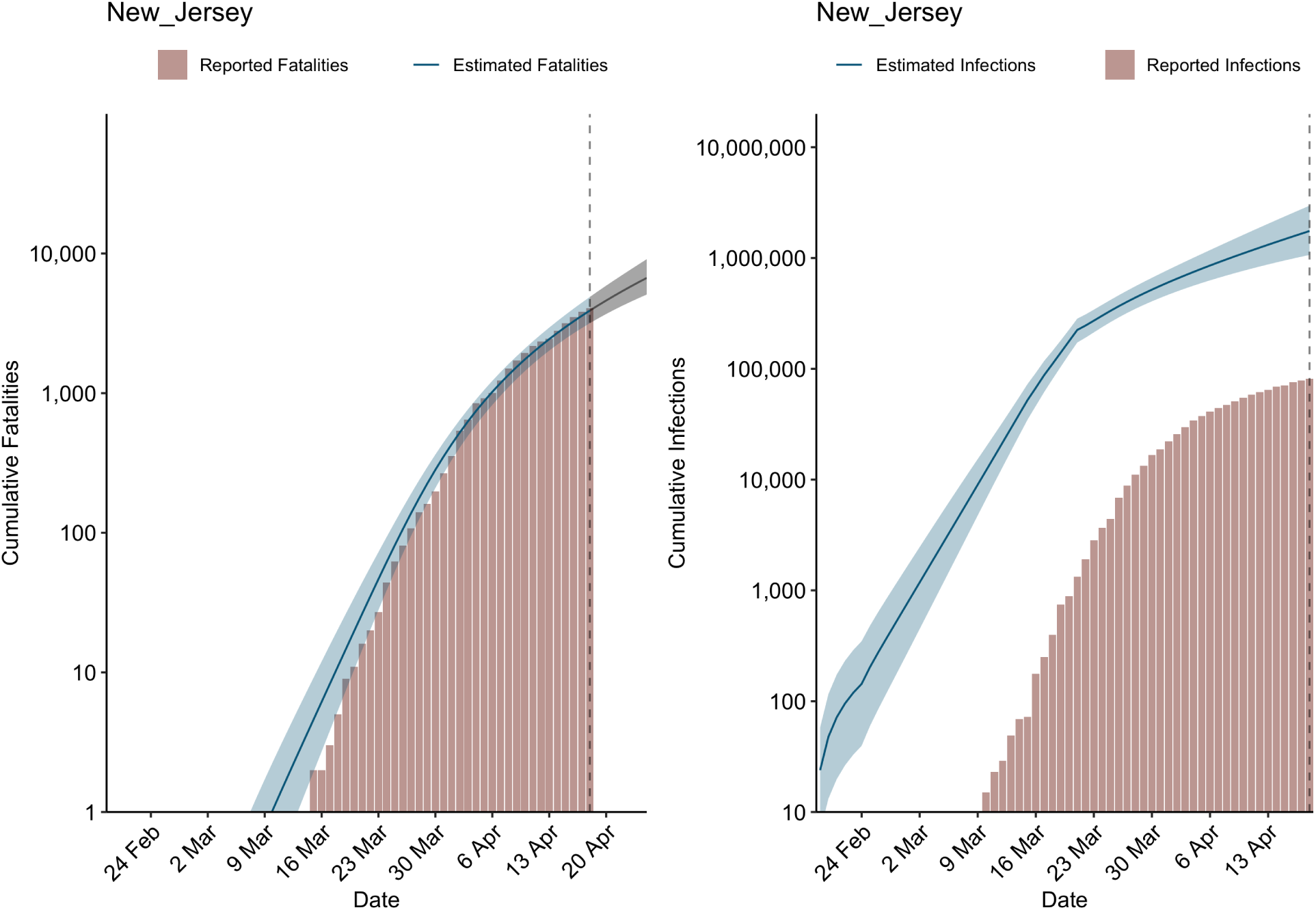
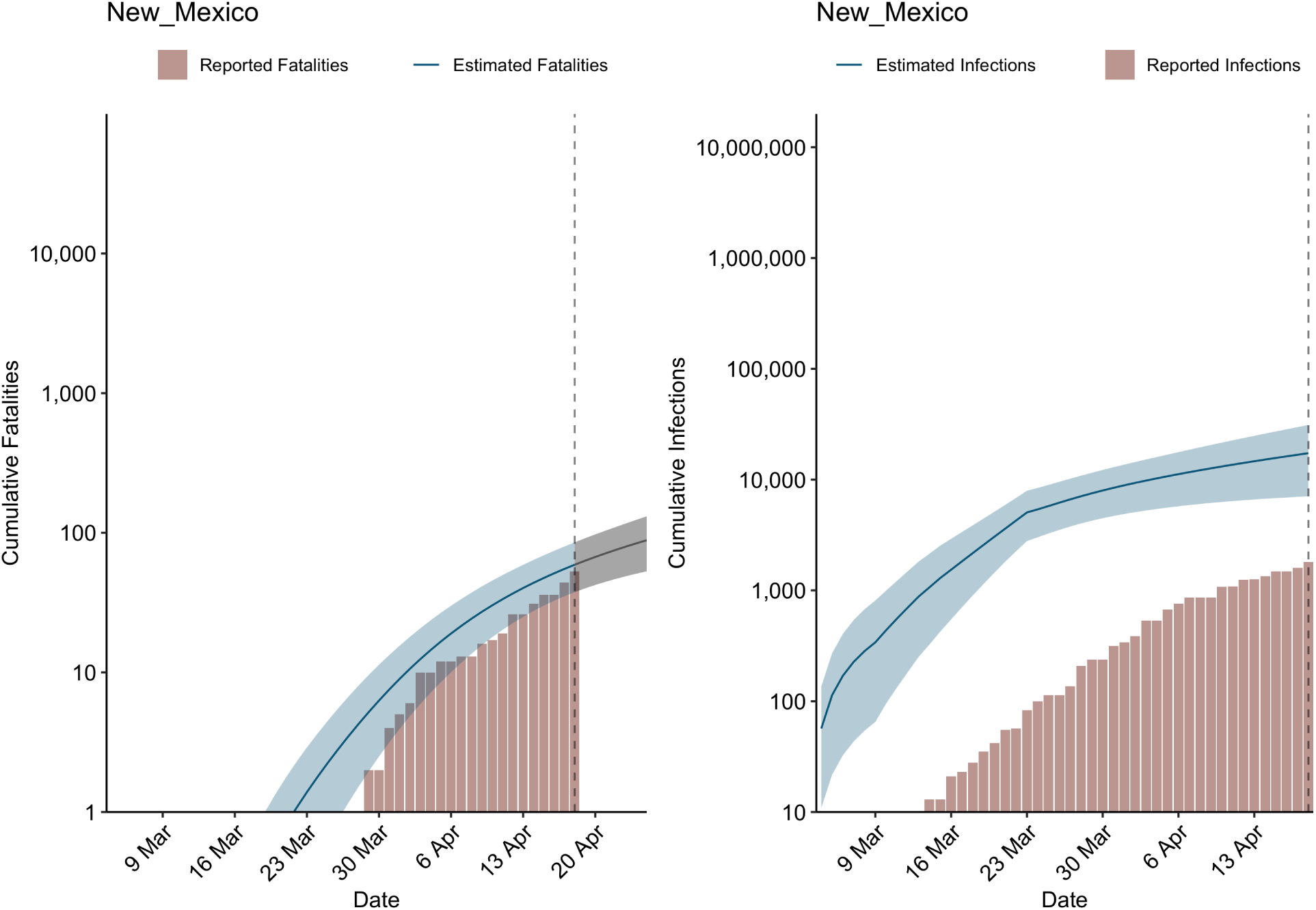
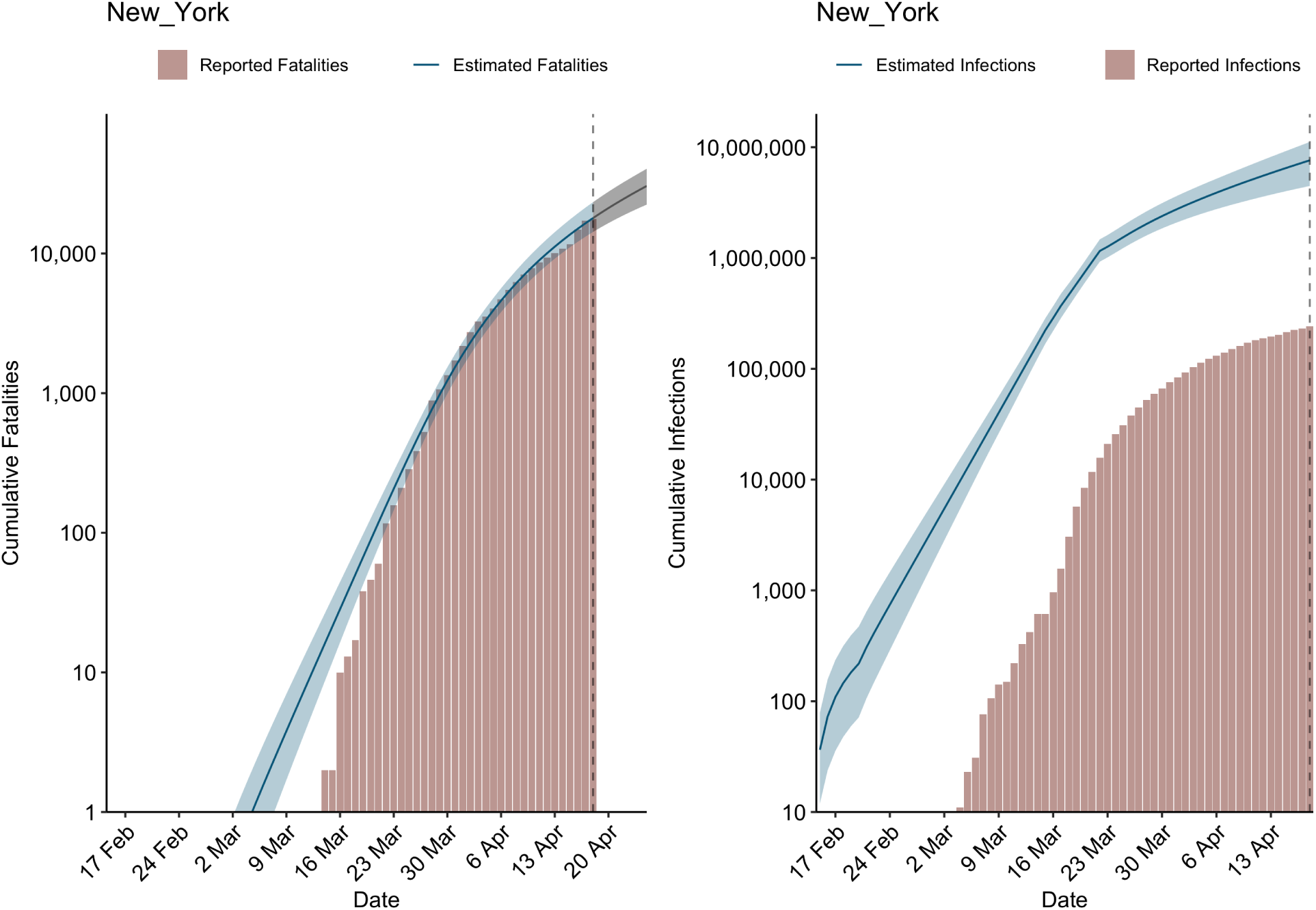
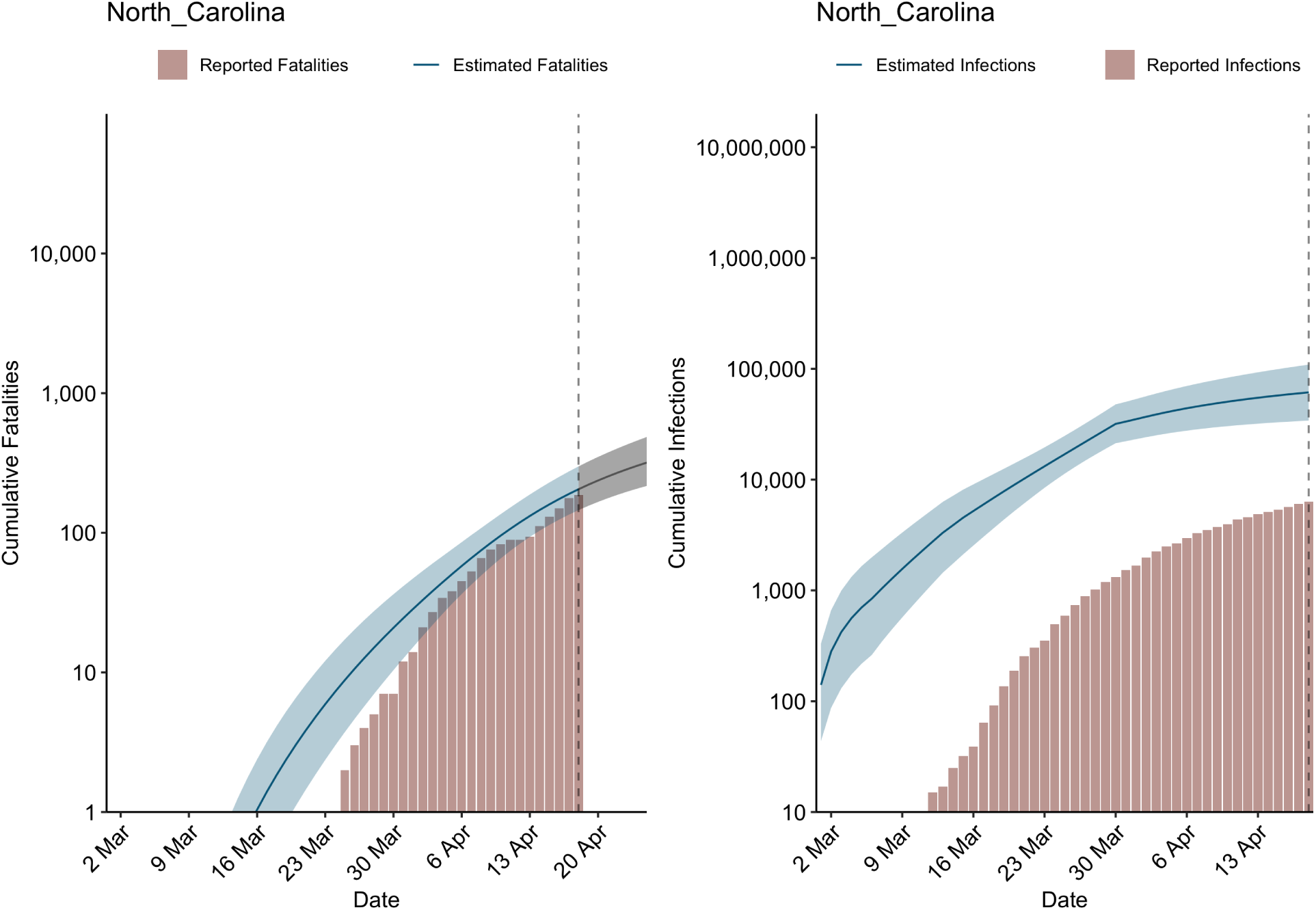
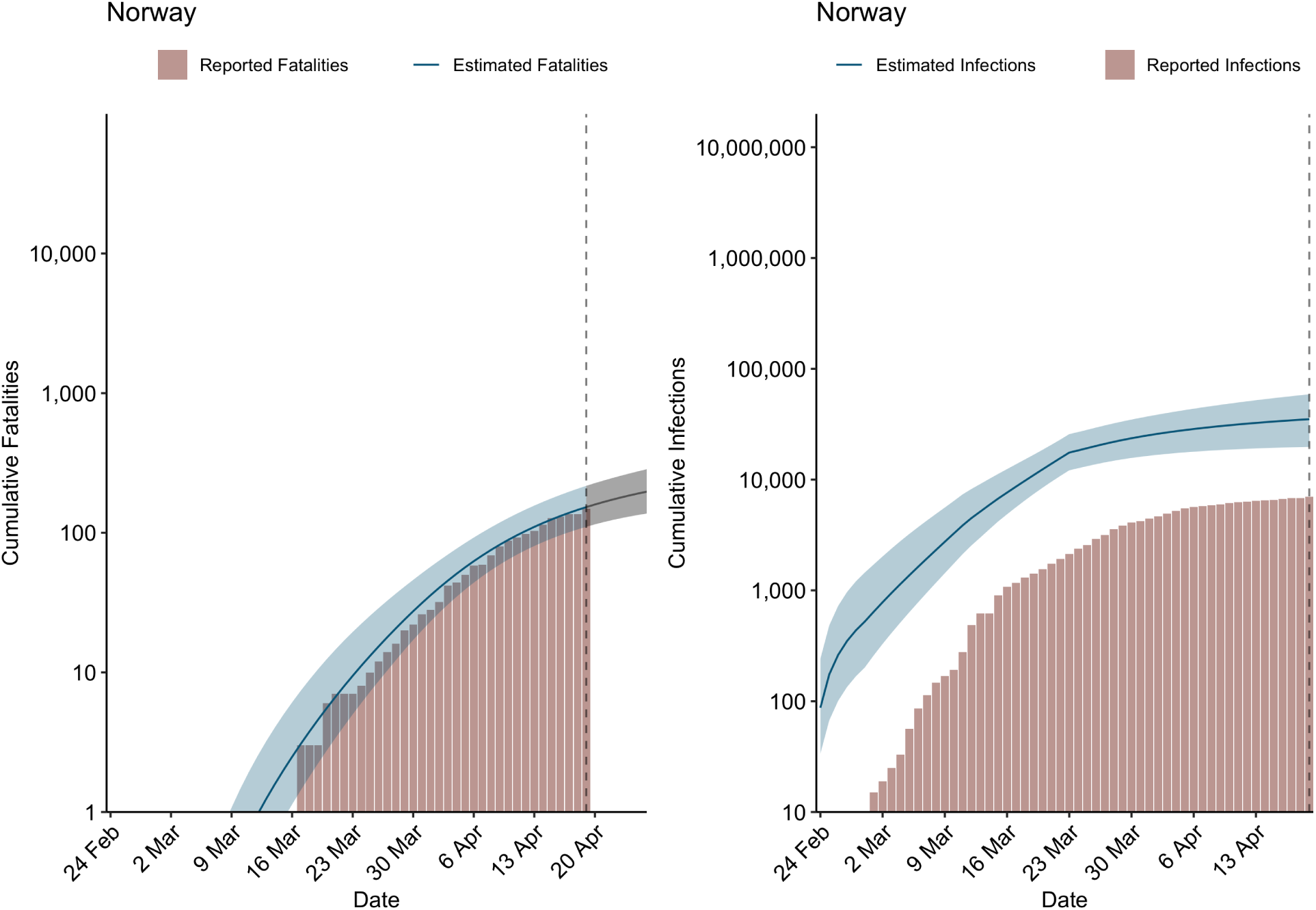
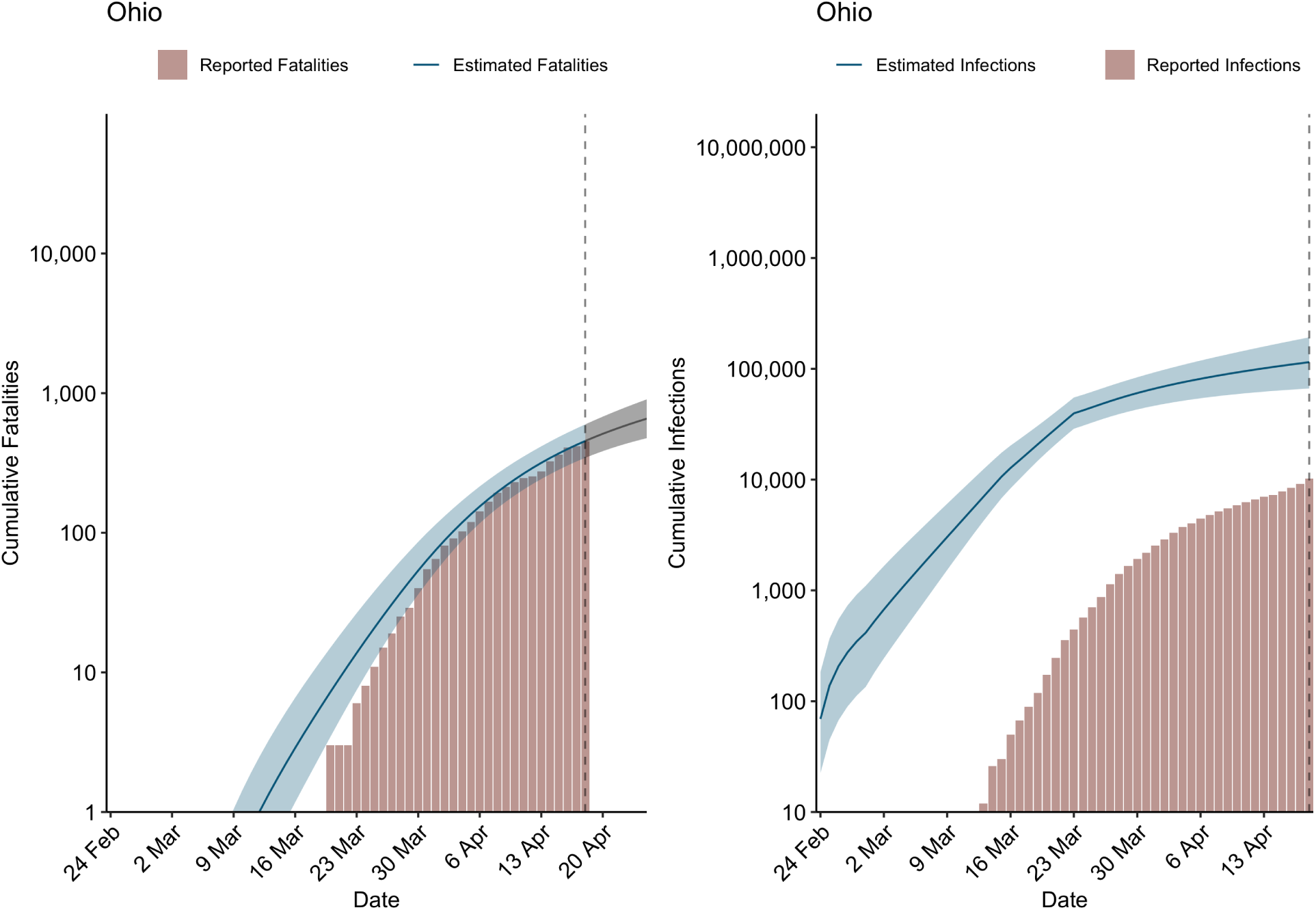
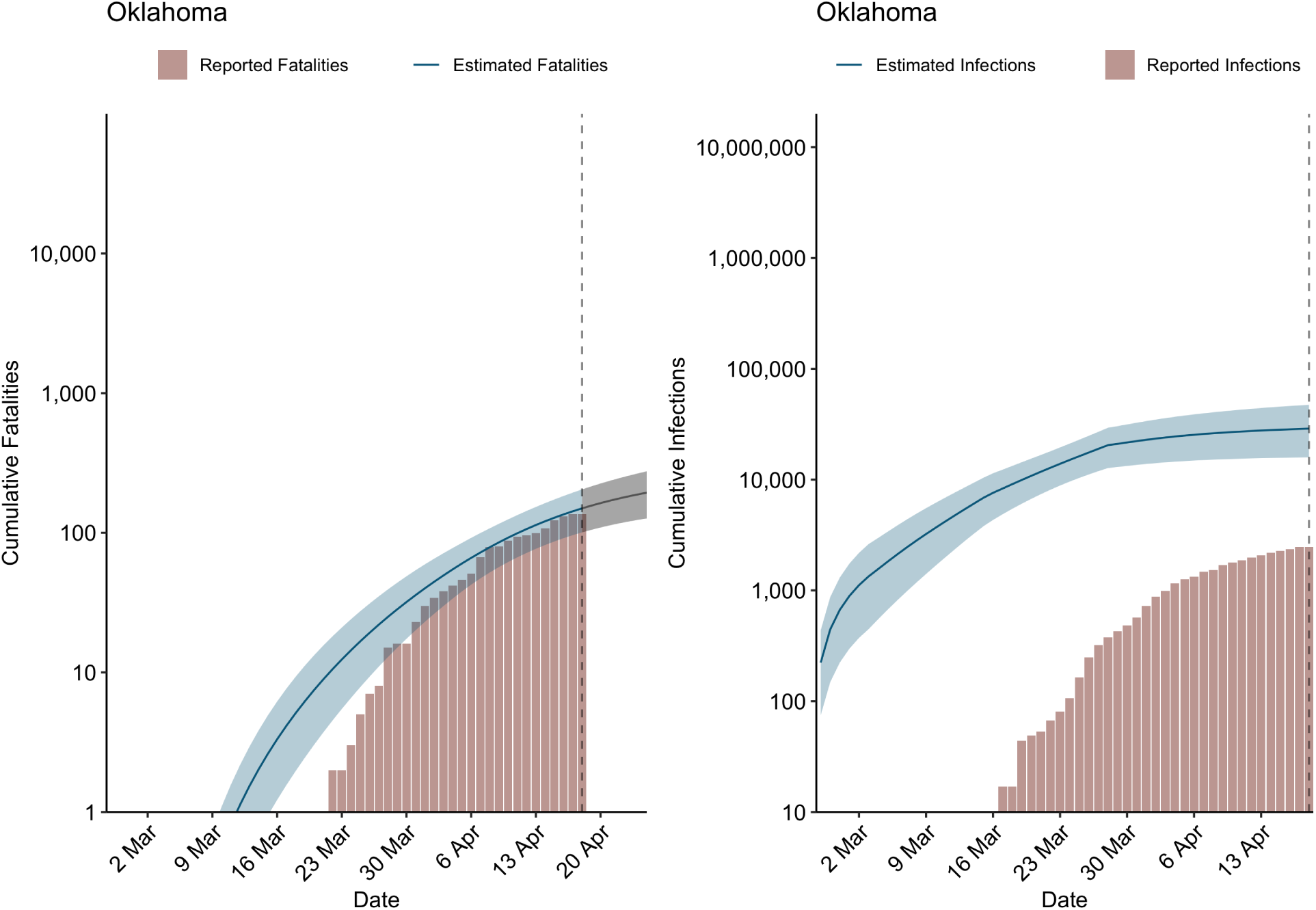
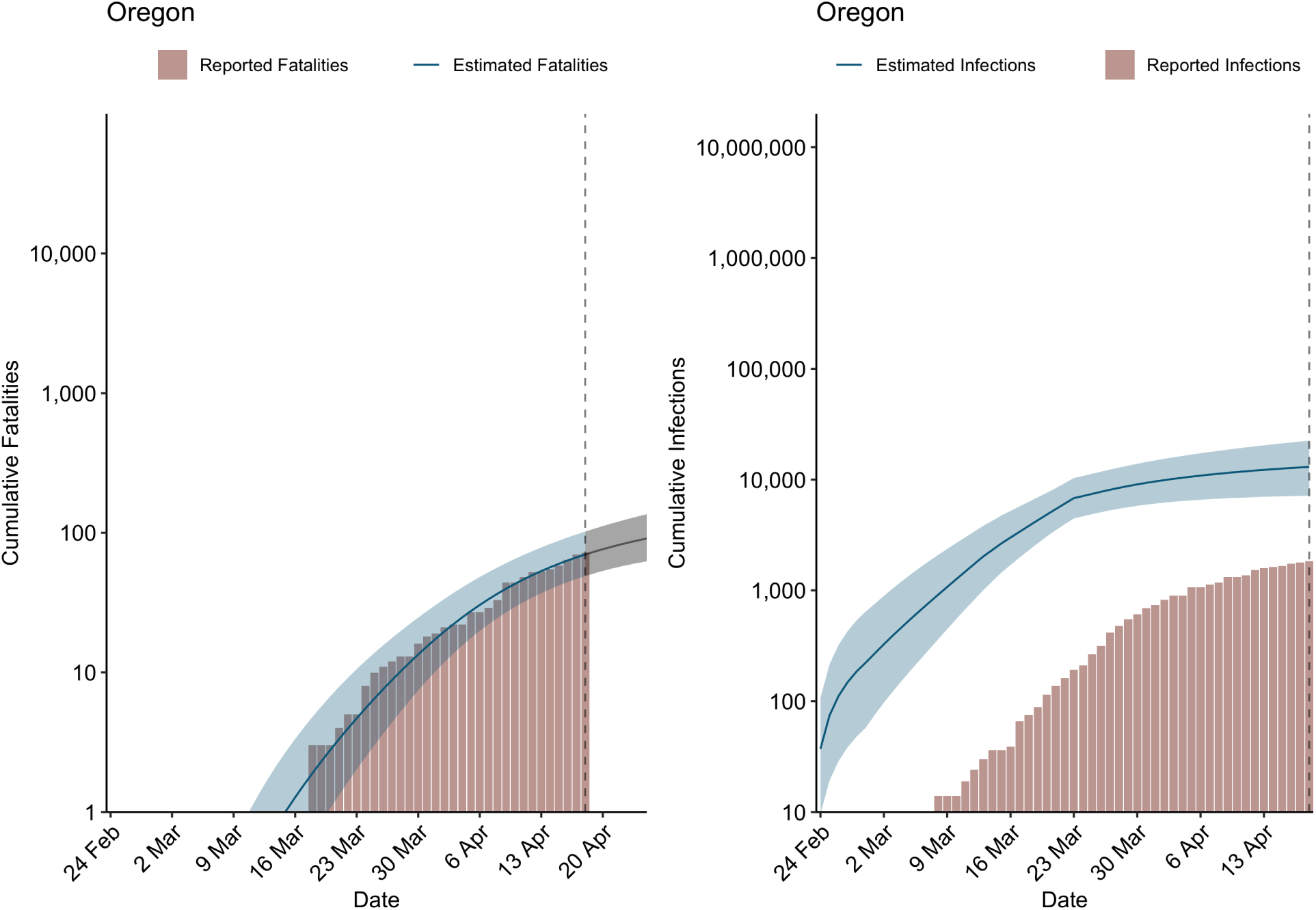
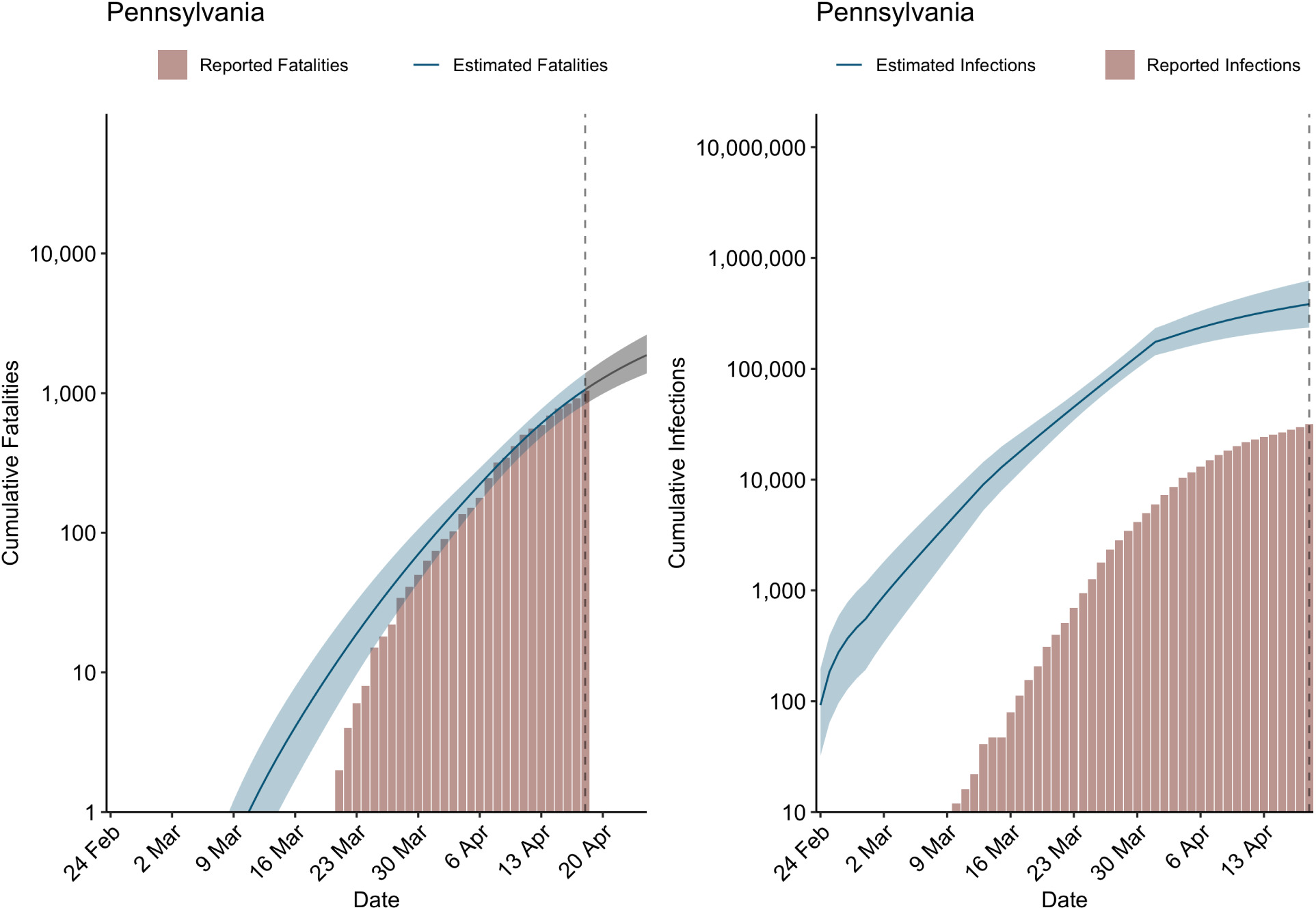
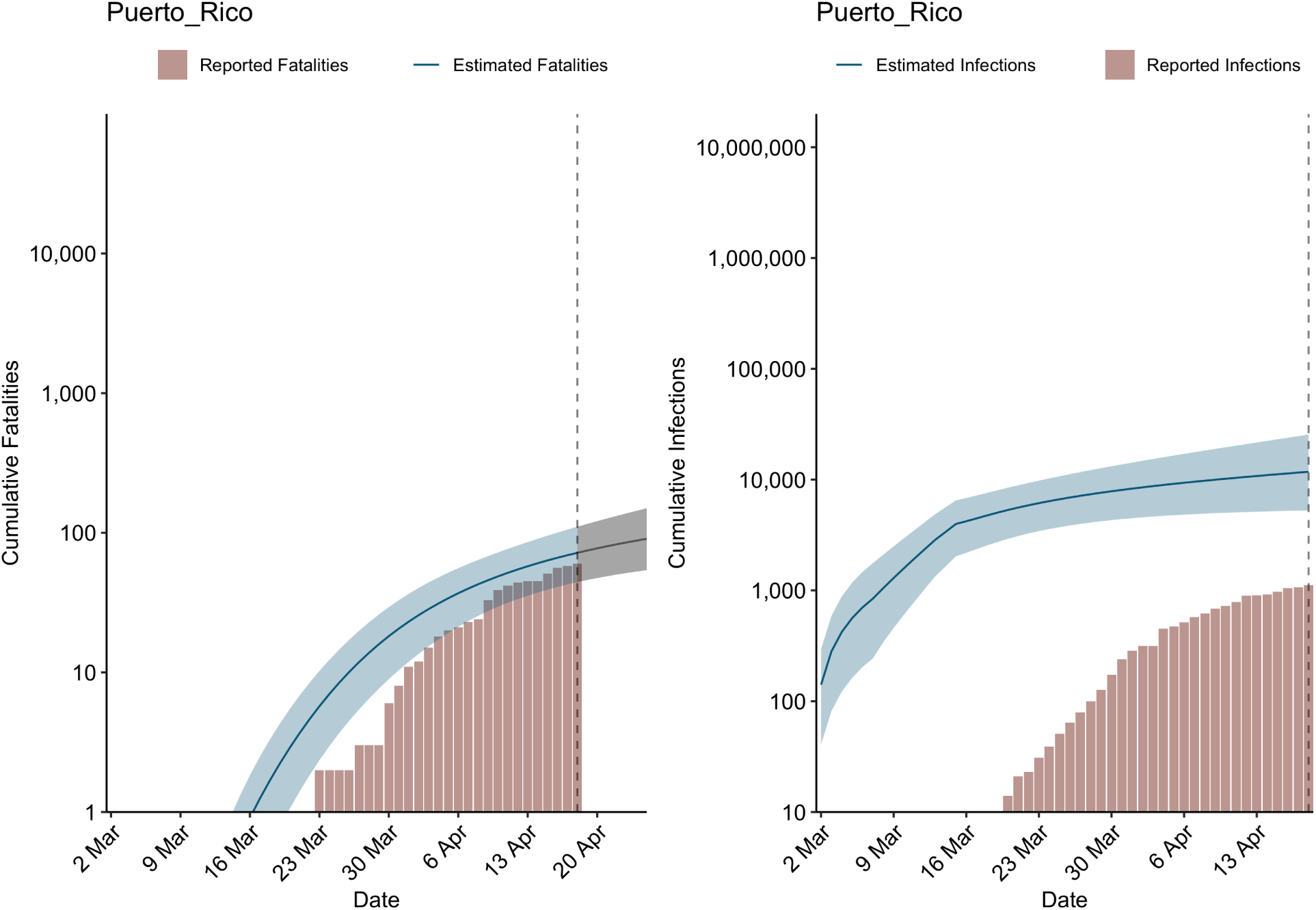
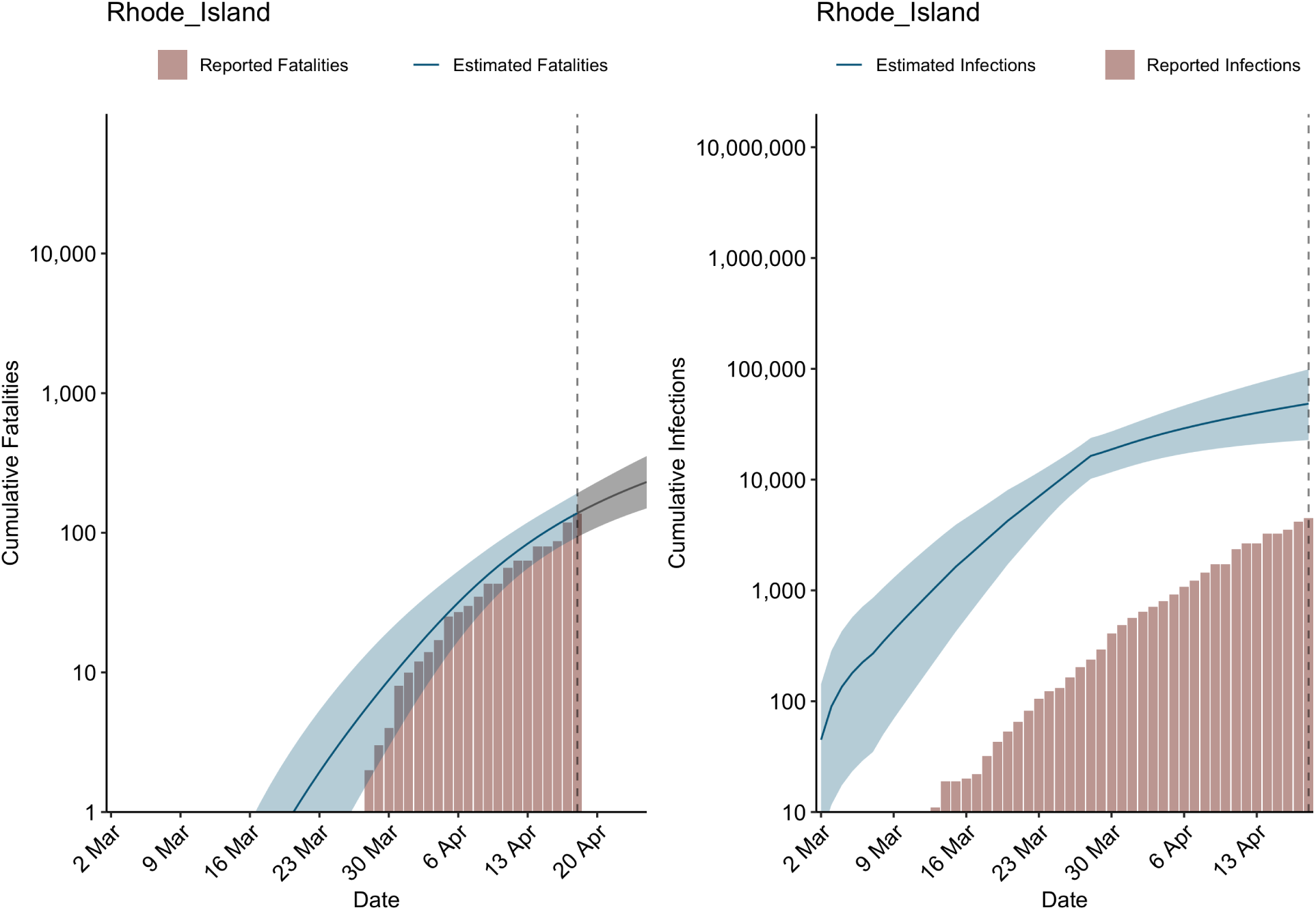
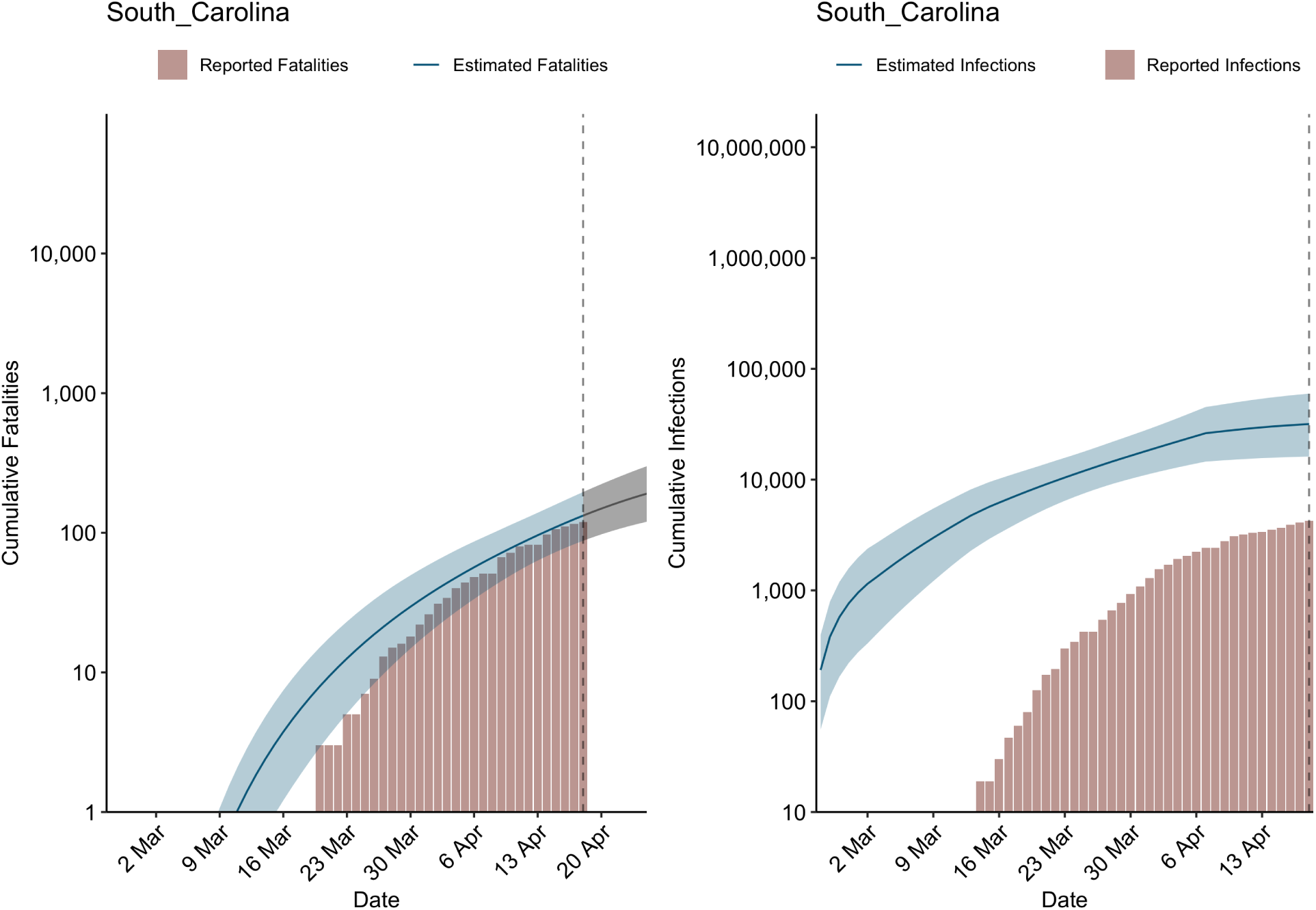
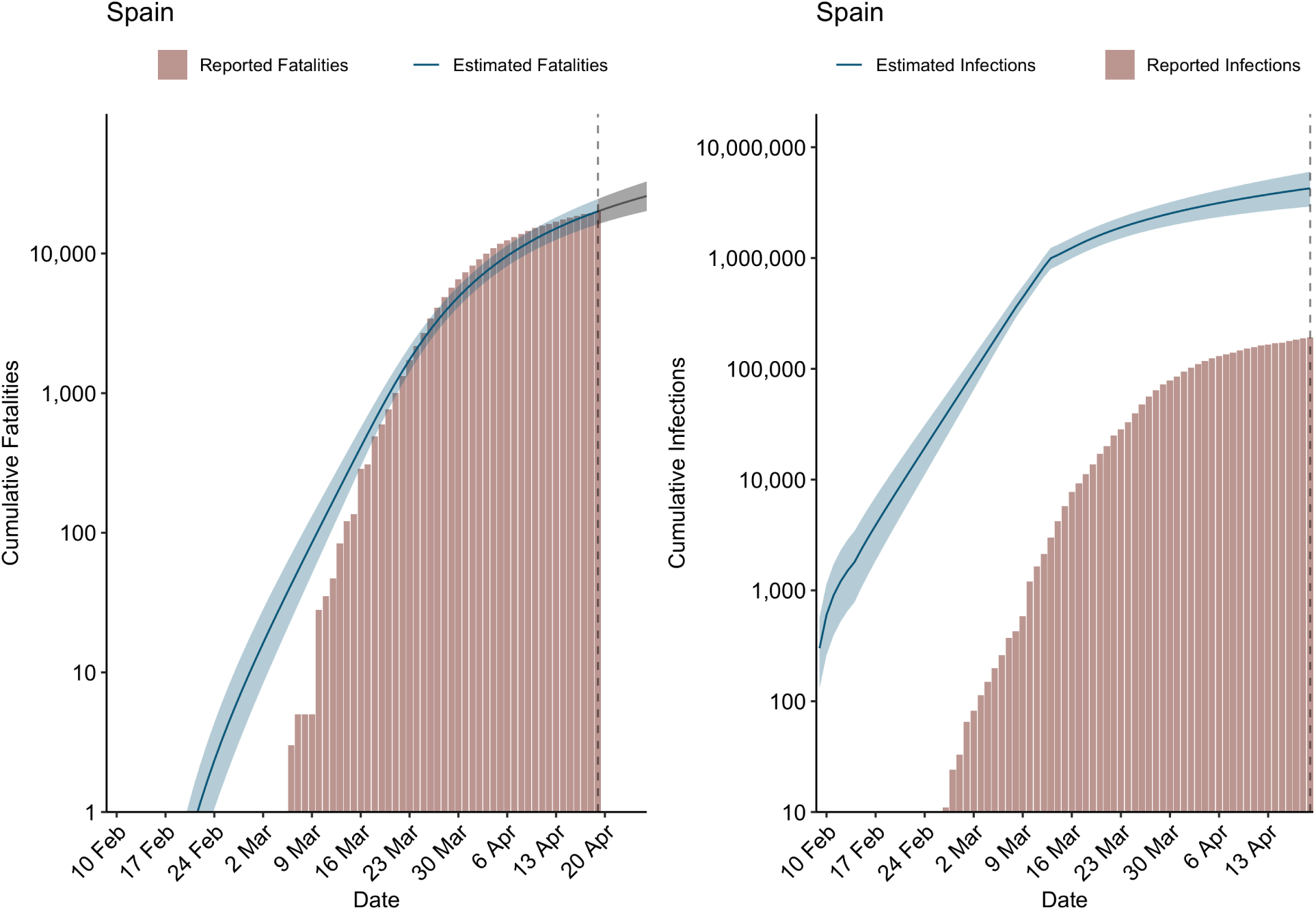
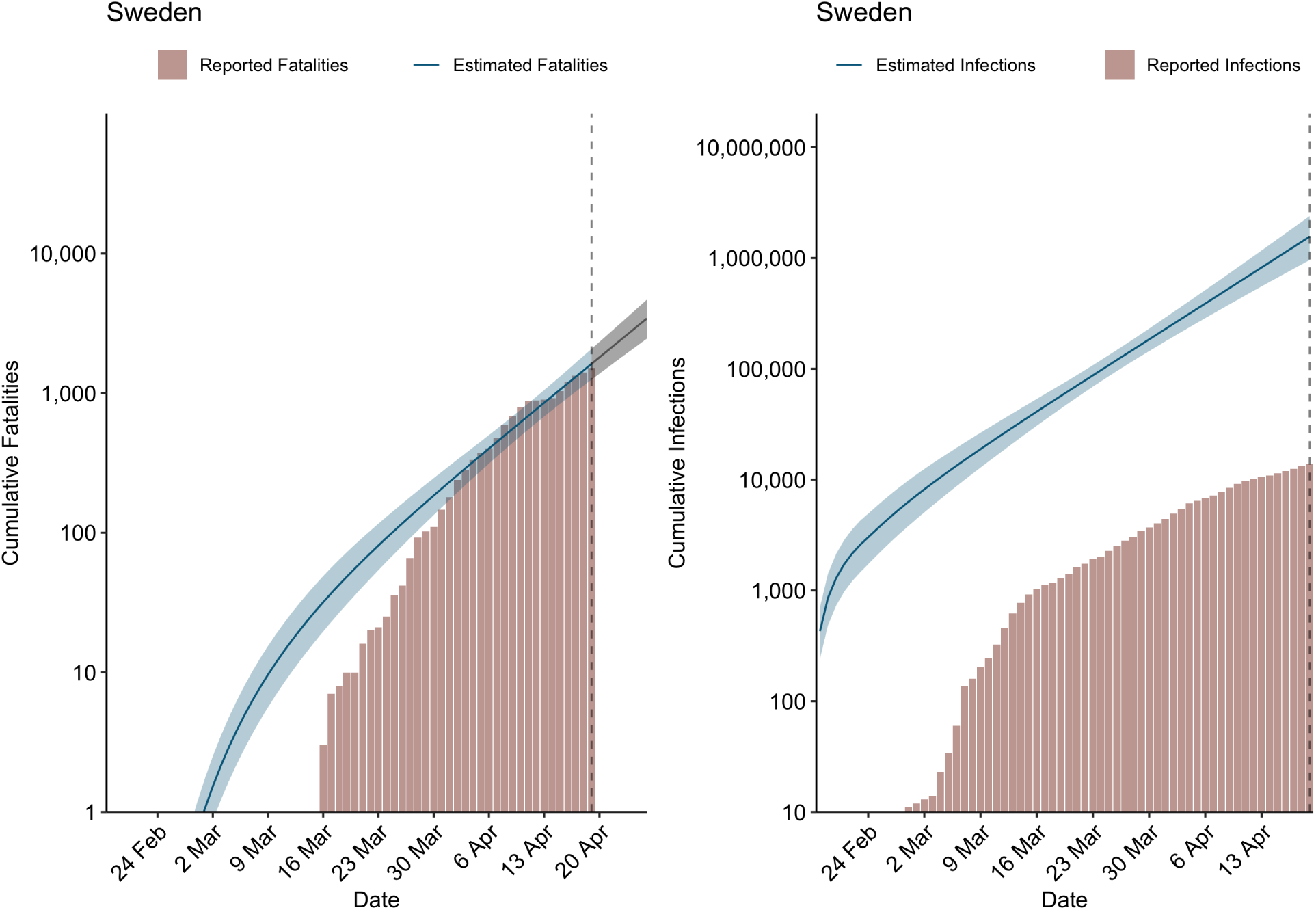
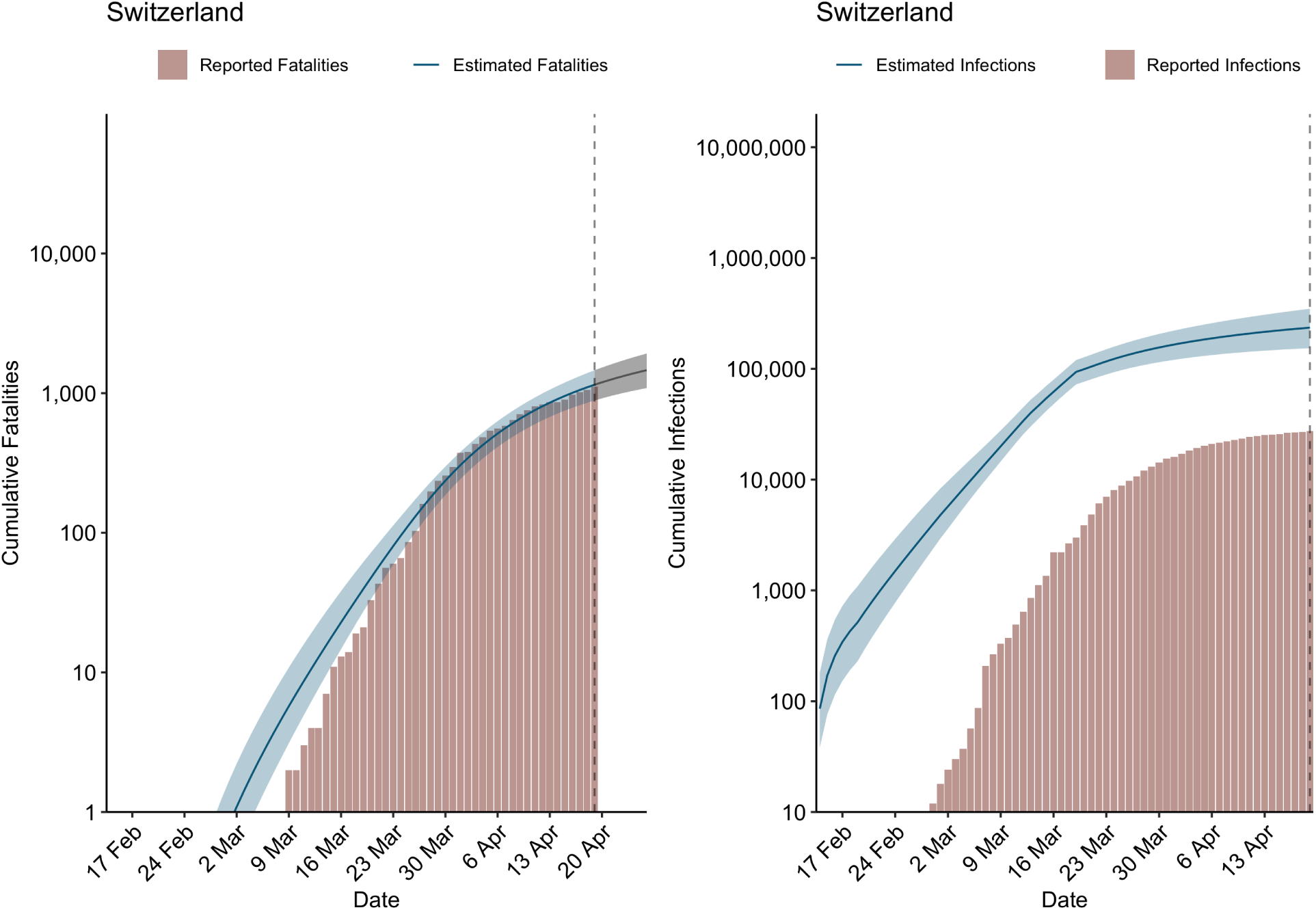
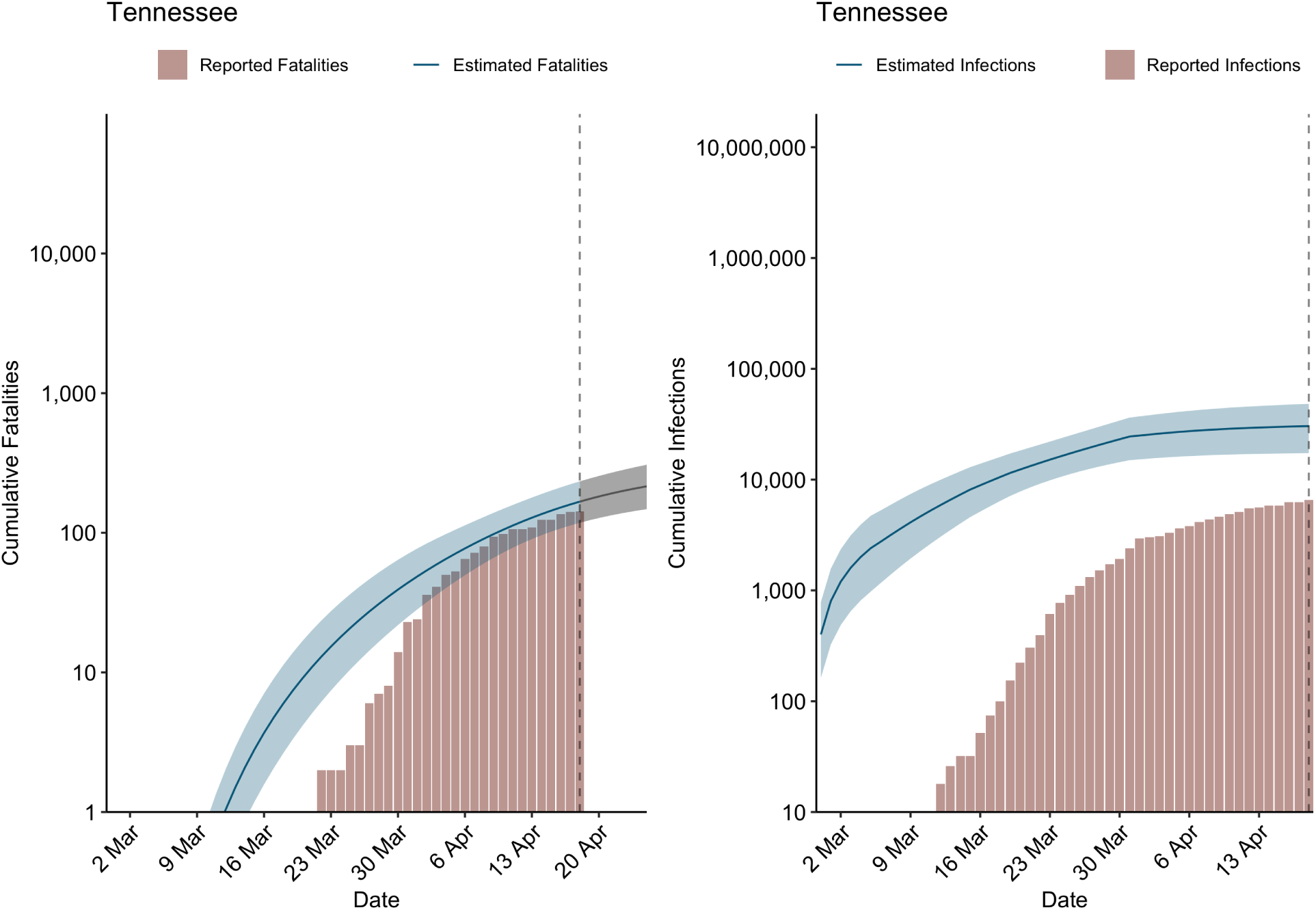
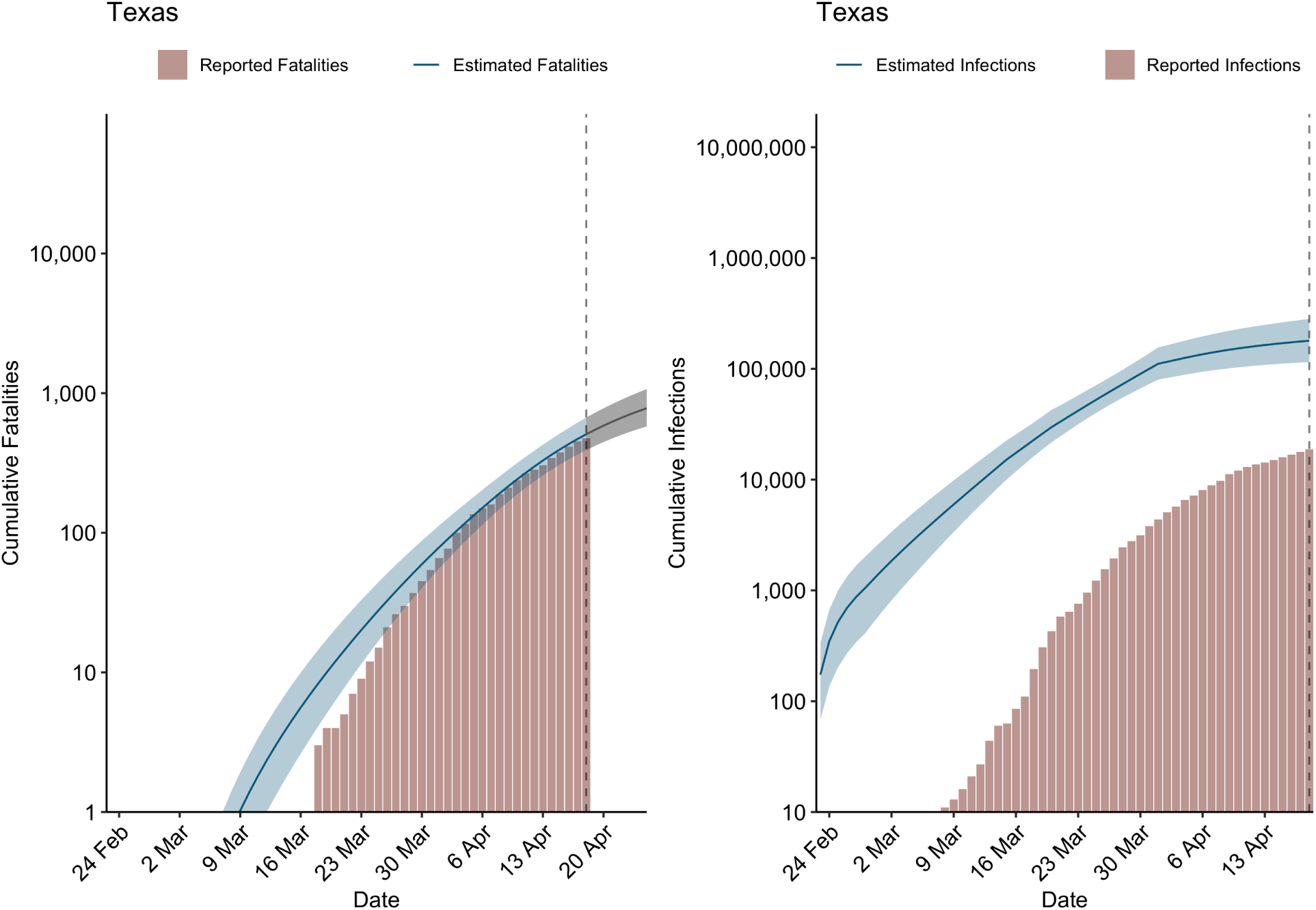
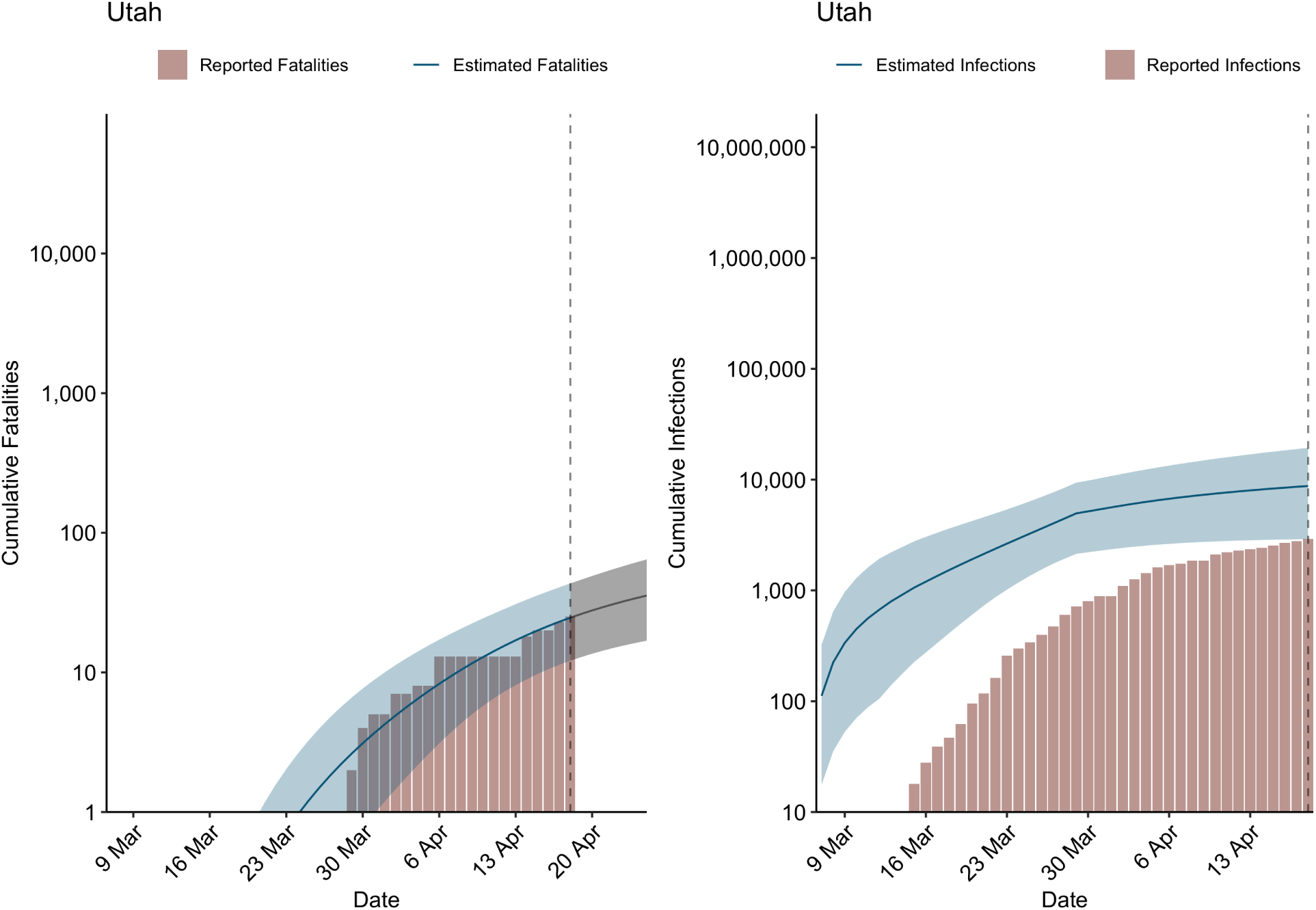
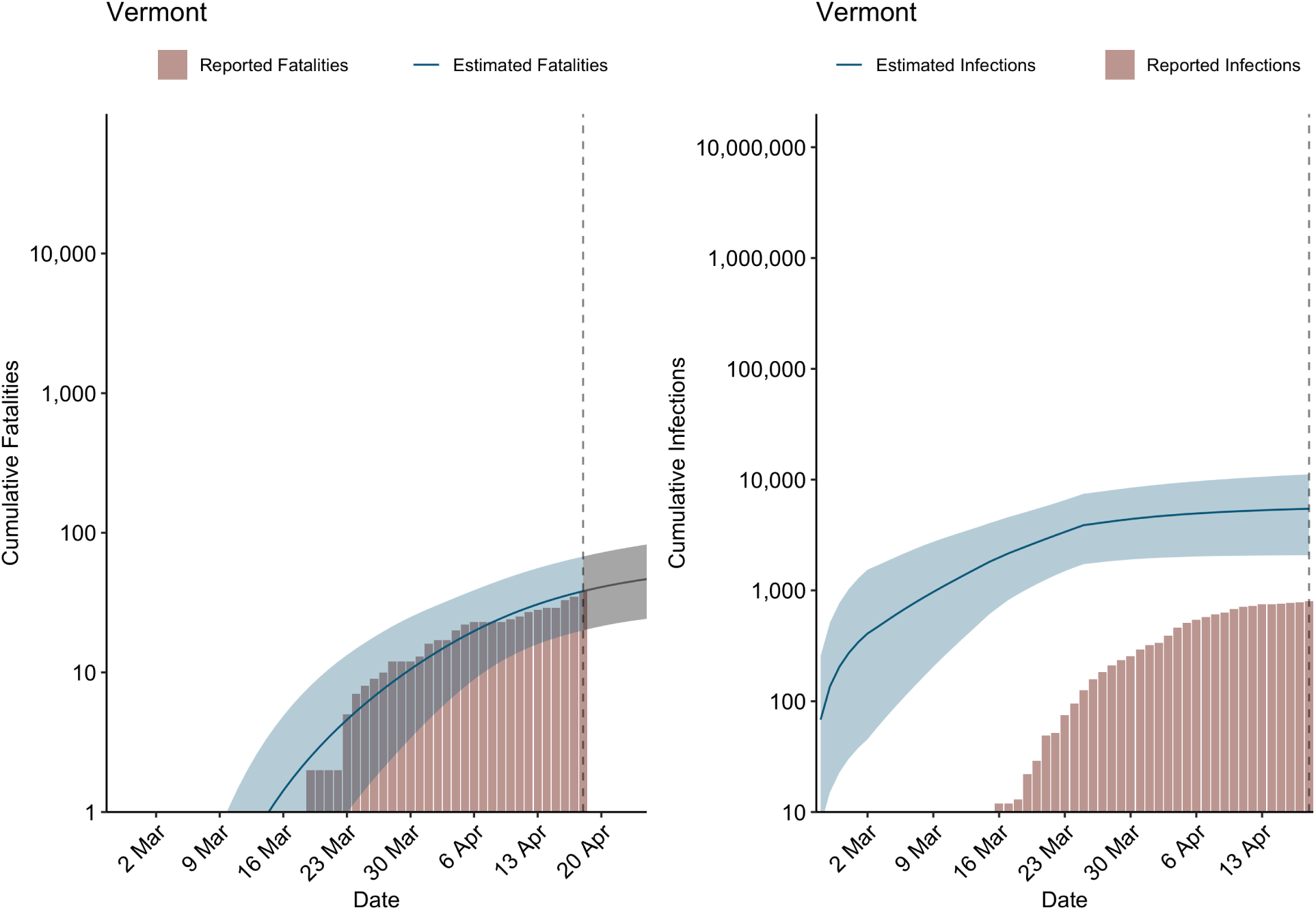
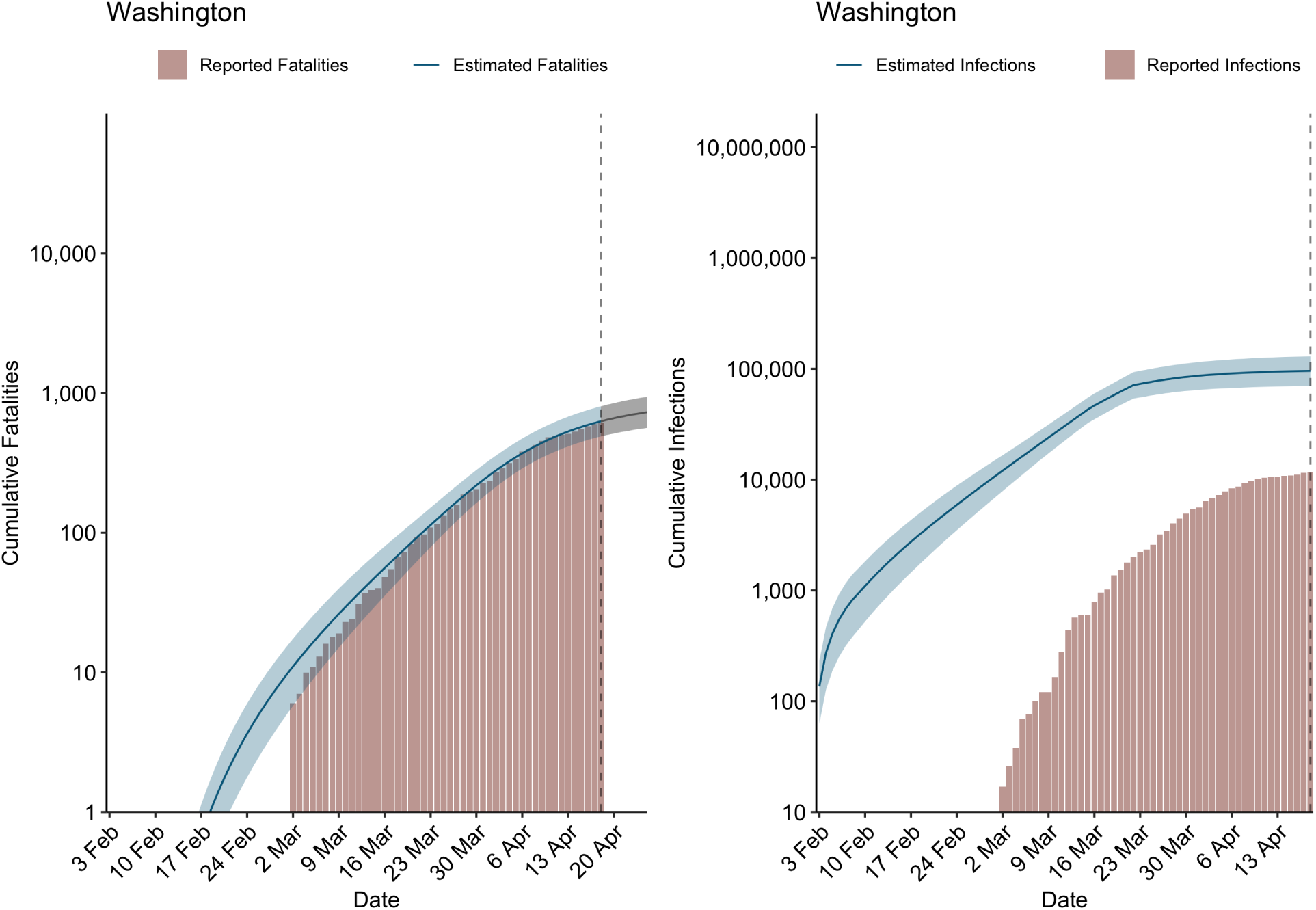
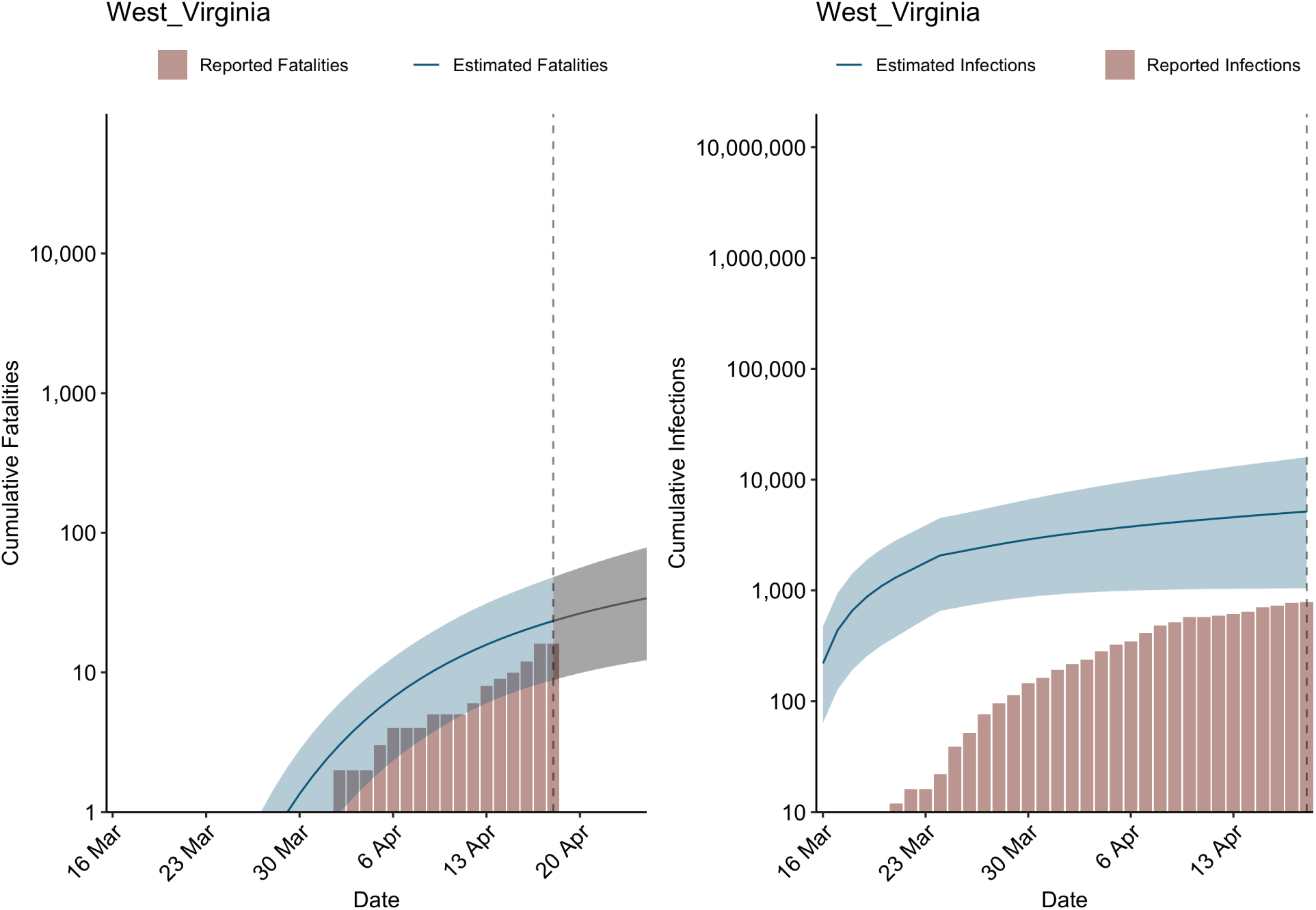
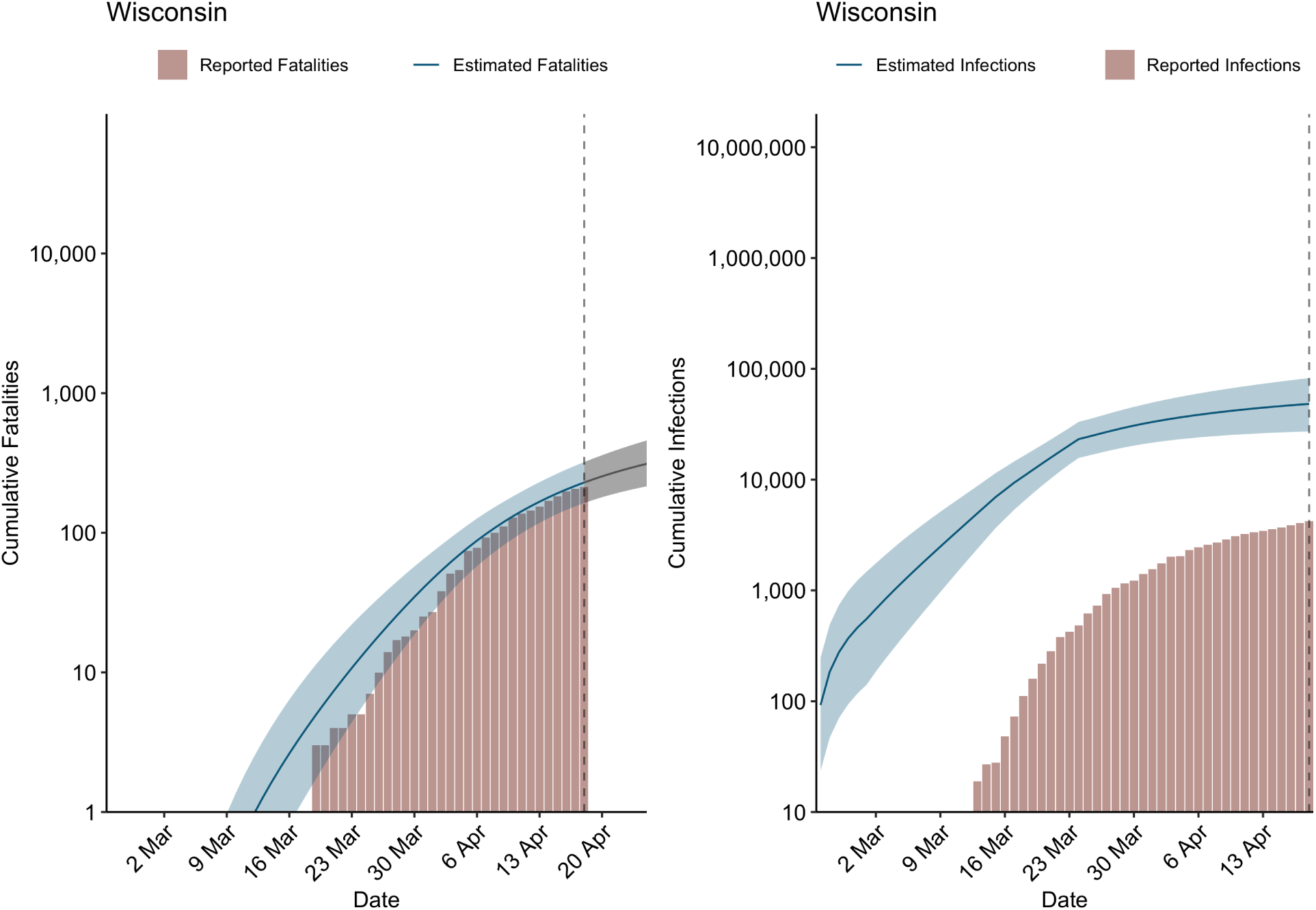

## Notes

### Competing Interest Statement

The authors have declared no competing interest.

### Funding Statement

We received no external funding for this work.

